# Membrane-Localized Mutations Predict the Efficacy of Cancer Immunotherapy

**DOI:** 10.1101/2022.05.28.22275728

**Authors:** Priscilla S. Briquez, Sylvie Hauert, Zoe Goldberger, Trevin Kurtanich, Aaron T. Alpar, Grégoire Repond, Yue Wang, Suzana Gomes, Prabha Siddarth, Melody A. Swartz, Jeffrey A. Hubbell

## Abstract

Due to their genetic instability, tumor cells bear mutations that can effectively be recognized by the immune system. In the clinic, immune checkpoint immunotherapy (ICI) can re-activate immune reactions against mutated proteins, known as neoantigens, leading to remarkable remission in cancer patients. Nevertheless, only a minority of patients are responsive to ICI, and approaches for prediction of responsiveness remain elusive yet are needed to improve the success of cancer treatments. While the tumor mutational burden (TMB) correlates positively with responsiveness and survival of patients undergoing ICI therapy, the influence of the subcellular localizations of the mutated proteins within the tumor cell has not been elucidated. Here, we hypothesized that the immune reactions are modulated by the localization of the mutated proteins and, therefore, that some subcellular localizations could favor responsiveness to ICI. We show in both a mouse melanoma model and human clinical datasets of 1722 ICI-treated patients that high membrane-localized tumor mutational burden (mTMB), particularly at the plasma membrane, correlate with responsiveness to ICI therapy and improved overall survival across multiple cancer types. We further highlight that mutations in the genes encoding for the membrane proteins *NOTCH3, RNF43, NTRK3* and *NOTCH1*, among others, may serve as potent biomarkers to predict extended survival upon ICI in certain cancer types. We anticipate that our results will improve the predictability of cancer patient response to ICI and therefore may have important implications to establish future clinical guidelines to direct the choice of treatment toward ICI.

## INTRODUCTION

Immunotherapies have revolutionized the landscape of clinical oncology, being established as first-line treatments in multiple advanced cancer types, including melanoma, non-small cell lung cancer (NSCLC) and renal cell carcinoma^1-3^. Despite the strong efficacy of immune checkpoint immunotherapy (ICI), less than 20% of patients show complete or durable response^4,5^. While studies have shown that infiltration of immune cells in the tumors^6^ and high tumor mutational burden (TMB) are key correlates of response to ICI^7-12^, accurate prediction of patient responsiveness to ICI remains an important challenge^13^. Greater predictivity certainly would increase patient survival and quality of life, by reducing the number, duration, and side-effects of treatments as well as associated economic burden.

Here, we hypothesized that the potency of immune response against tumor mutated proteins not only depends on the total mutational burden, but also on the subcellular localization of these proteins within the tumor cell. Indeed, the efficiency of presentation of mutated proteins on the major histocompatibility complex (MHC)-I by the tumor cell, required for recognition and killing by CD8^+^ T cells^14^, might vary for cytoplasmic, nuclear, membrane-localized or secreted proteins due to their specific intracellular processing and trafficking routes^15-17^. In addition, efficiency in collection and presentation of these mutated proteins by antigen-presenting cells (APCs), on both MHC-I and -II to generate CD8^+^ and CD4^+^ T cell responses, respectively, could similarly be impacted by these different forms of proteins upon release in debris, vesicles or in the extracellular milieu. Apart from antigen presentation, membrane-bound mutated proteins can be recognized by antibodies, induced via B cell immunity, which could allow antibody-dependent cytotoxic mechanisms that kill tumor cells by activating natural killer (NK) cells, macrophages or the immune complement cascade^18,19^. Of note, tumor mutated proteins that successfully activate an adaptative immune response are commonly defined as tumor neoantigens.

To date, very few reports have examined how the subcellular localization of tumor mutated proteins modulates anti-cancer immunity. In this study, we show that high membrane-localized tumor mutational burden (mTMB) increase tumor immunogenicity and improve responsiveness to ICI therapies. We first demonstrated in a mouse model of melanoma that membrane-localization of OVA (mOVA; used here as a model tumor antigen) in B16-F10 cells increased local and systemic immunity as compared to soluble OVA and rendered these tumors highly susceptible to ICI, in a manner that did not depend on immunoglobulin G (IgG) antibody-mediated cytotoxicity. We then questioned if a high mTMB improves responsiveness to ICI in cancer patients. We developed a simple algorithm that extracts the subcellular localizations associated with tumor mutated genes from the UniprotKB/Swiss-Prot database^20^ and analyzed the publicly available sequencing data of 4864 patients, treated or not with ICI, from studies by Samstein *et al*.^7^, Hellman *et al*.^8^ and Hugo *et al*.^9^. We demonstrated that high mTMB correlates with increased patient survival and responsiveness to ICI across multiple cancer types. Moreover, we highlighted that mutated genes encoding for some particular membrane-localized proteins may serve as potent biomarkers to predict extended survival of patients upon ICI, such as *NOTCH1, NOTCH3, RNF43* or *NTRK3*. Together, our results highlight the importance of considering the subcellular localization of tumor mutated proteins, in particular mTMB, in addition to the total TMB, to improve the predictivity of patient responsiveness to ICI therapy and potentially the clinical guidelines for the selection of the most appropriate cancer treatment. Such findings may also have strong implications on vaccinal antigen selection for neoantigen-targeted cancer vaccines based on tumor gene sequencing.

## RESULTS

### Membrane-bound antigens increase tumor immunogenicity

We began by studying the effect of cell membrane-bound antigens in the B16-F10 murine melanoma model. We first modified B16-F10 cells for expression of membrane-bound OVA (B16mOVA), by fusing the full-length OVA sequence to the transmembrane domain of H-2D^B^ (Fig. S1a)^21^. As a control, we used B16-F10 cells that expresses full-length OVA in a soluble form (i.e., not membrane-bound; B16-OVA). For both designs, we generated cell lines with matching high (^HI^) and low (^LO^) levels of OVA expression, as quantified by qPCR (Fig. S1b, c). The presence of OVA at the surface of the B16mOVA cells, but not on the B16-OVA cells, was confirmed by flow cytometry and fluorescence (Fig. S1d, e). We further highlighted that membrane-bound OVA was secreted on extracellular vesicles produced by B16mOVA (Fig. S1f), which is potentially important to increase antigen transport and availability to APCs.

Upon intradermal injection in C57BL6 wild-type (WT) mice, all cell lines were tumorigenic. We observed that B16mOVA^HI^ tumors grew significantly slower than B16-OVA^HI^ and the parental B16 WT, which resulted in extended survival of mice bearing B16mOVA^HI^ tumors (Fig. 1a, Fig. S1g). This effect was antigen dose-dependent, as seen by an intermediate growth rate of the B16mOVA^LO^ tumors. To confirm that this difference was due to immune-mediated rejection of the tumor, rather than to a difference in cell growth/division rate, we evaluated B16mOVA^HI^ tumor growth in transgenic Act-mOVA mice, which are immune tolerant to mOVA. In these mice, B16mOVA^HI^ tumors grew faster than the B16 WT tumors, demonstrating an intact proliferation capacity of the B16mOVA^HI^ cells (Fig. S1h). This supports the hypothesis that the slowed tumor growth in WT mice was due to an immune reaction against mOVA.

Therefore, we analyzed immune cell infiltrates in the different OVA-expressing B16-F10 tumors, reasoning that increased OVA-mediated tumor rejection would enhance the local presence of inflammatory cells (Fig. S2, S3). Indeed, we found a significant increase of CD45^+^ immune cells in tumors that expressed mOVA as compared to dose-matched soluble OVA, of about 2-fold in the case of B16mOVA^HI^ vs. B16-OVA^HI^ tumors (Fig. 1b). Particularly, CD8^+^ T cells and NK cells were more numerous in this tumor, but not CD4^+^ T cells (Fig. 1b, Fig. S4a-c). No difference in PD-1 expression was observed on the T cells in the tumors expressing mOVA versus soluble OVA (Fig. S4d). Among the other immune cell types screened, NKT cells were slightly increased, and dendritic cells and B cells slightly decreased in the B16mOVA tumors when quantified relative to the total CD45^+^ immune cell population (Fig. S4e).

**Figure 1.**
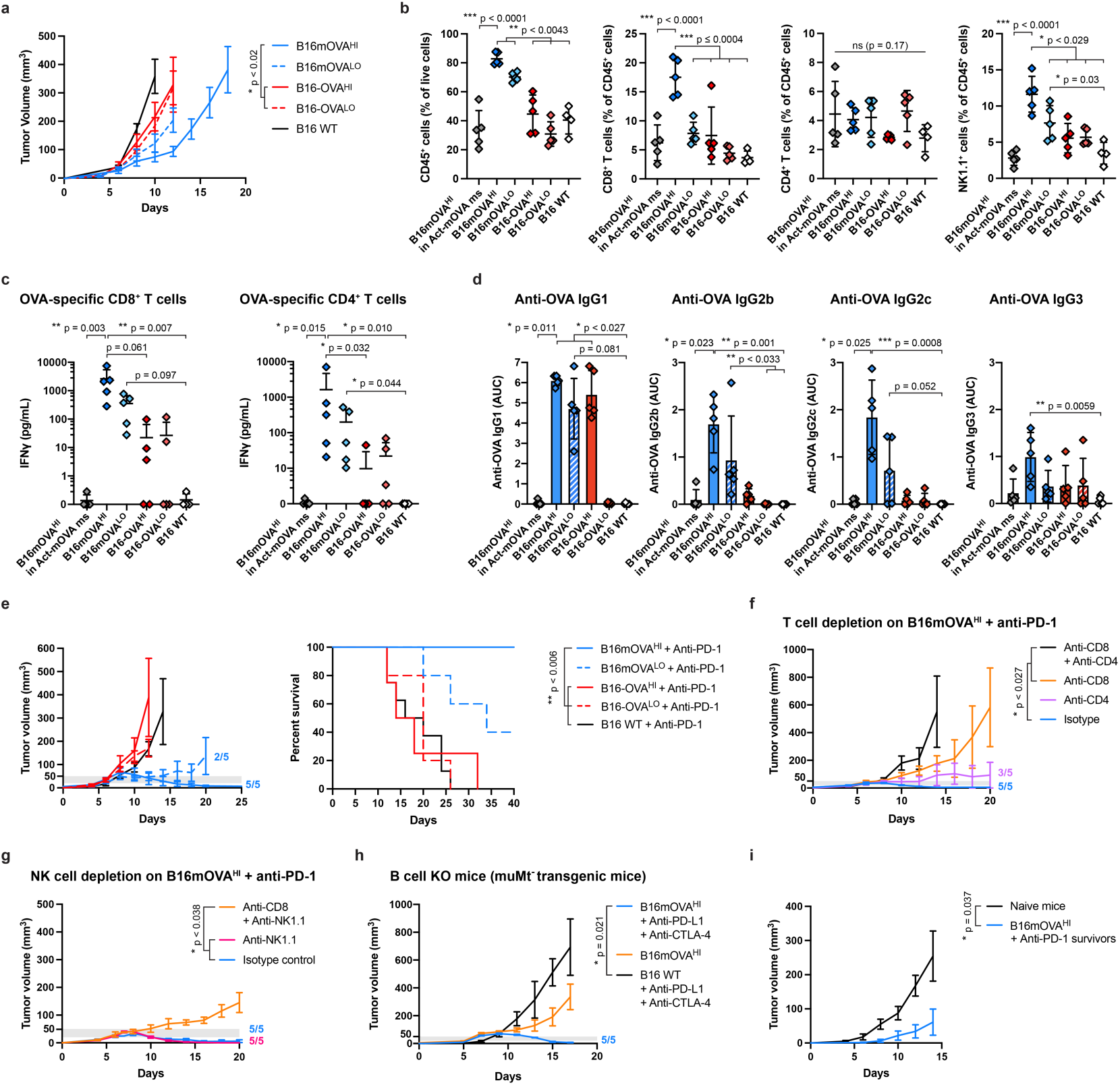
Membrane-bound antigens enhance melanoma tumor immunogenicity and responsiveness to ICI in mice. B16-F10 melanoma cells (1.5 M) modified to express membrane-bound or soluble full-length ovalbumin (B16mOVA and B16-OVA, respectively), at high (^HI^) or low (^LO^) levels, were injected intradermally in C57BL6 mice. The parental B16-F10 wild-type (WT) cells were used as a control. Where indicated, treatment with 200 μg of anti-PD-1 injected intraperitoneally was given to mice when their tumor volume reached 20-50 mm^3^ (grey thresholds). **a**, Tumor growth of the different OVA-expressing B16 cell lines upon injection *in vivo* (N≥8, mean ± SEM, Kruskal-Wallis with Dunn’s post-test at day 12). **b**, Immune cell populations infiltrated in the different tumors at day 10 post-injection analysed by flow cytometry (N≥4, mean ± SD, ANOVA with Tukey’s post-test and Brown-Forsythe correction when needed). **c**, *Ex vivo* restimulation of OVA-specific CD8^+^ and CD4^+^ T cells in spleen of mice bearing the different OVA-expressing tumors at day 10 post-injection (N≥4, mean ± SD, Kruskal-Wallis with Dunn’s post-test). **d**, Anti-OVA antibody quantification per IgG subtype in the plasma of tumor-bearing mice at day 10 post-injection (AUC: area under the curve; N≥4, mean ± SD, Kruskal-Wallis with Dunn’s post-test). **e**, Tumor growth and associated survival of OVA-expressing tumor-bearing mice treated with anti-PD-1 (N≥5, mean ± SEM, log-rank tests with Holm-Bonferroni p-values adjustment). **f**, B16mOVA^HI^ tumor growth upon depletion of CD8^+^ or/and CD4^+^ T cells with anti-PD1 treatment (N≥5, mean ± SEM, Kruskal-Wallis with Dunn’s post-test at day 14). **g**, B16mOVA^HI^ tumor growth upon depletion of NK1.1^+^ or/and CD8^+^ T cells with treatment with anti-PD1 (N≥5, mean ± SEM, Kruskal-Wallis with Dunn’s post-test at day 20). **h**, B16mOVA^HI^ tumor growth in MuMt^-^ mice (lacking mature B cells) with treatment with anti-PD-1 (N≥4, mean ± SEM, Kruskal-Wallis with Dunn’s post-test at day 17). **i**, Tumor growth of mice that survived B16mOVA^HI^ tumors treated with anti-PD-1 upon rechallenge with 250k B16-F10 WT cells (N≥8, mean ± SEM, Mann-Whitney test at day 14).

Next, we assessed whether immunity against OVA in the B16mOVA-bearing mice was sufficiently strong to induce systemic immunity, in addition to local intratumoral inflammation. *Ex vivo* restimulation of splenocytes using OVA-derived MHC-I and MHC-II peptides revealed that both CD8^+^ and CD4^+^ T cell responses were increased in mice with tumor expressing mOVA as compared to those with soluble OVA, as highlighted by the production of the pro-inflammatory cytokine interferon (IFN)-γ (Fig. 1c). In addition, OVA-specific antibody responses were detected in the plasma of tumor-bearing mice for both B16mOVA and B16-OVA, but different subtypes of immunoglobulin G (IgG) were generated, depending on the antigen localization. Particularly, OVA-specific IgG2b and IgG2c were detected in mice bearing B16mOVA tumors but were largely absent in those bearing B16-OVA tumors (Fig. 1d, Fig. S4f).

Together, these results showed that membrane-bound tumor antigens, here modelled by mOVA, strongly enhanced tumor immunogenicity both locally and systemically, resulting in slowed tumor growth and extended survival of untreated mice.

### Membrane-bound antigens restore responsiveness to ICI

While B16-F10 WT melanoma does not respond ICI, we examined whether the increased immunogenicity of the B16mOVA, particularly the enhanced presence of intratumoral T cells, would render them more susceptible. Remarkably, all mice (5 out of 5) bearing B16mOVA^HI^ tumors and treated with anti-PD1 therapy showed complete responses to ICI, whereas B16-OVA^HI^ and B16 WT-bearing mice were completely unresponsive (Fig. 1e). Lowering the antigen dose in the B16mOVA^LO^ group reduced the efficacy of ICI yet resulted in 2 out of 5 tumor eradications and otherwise slowed tumor growth. Such effects were also confirmed using the combination therapy anti-PD-L1 and anti-CTLA-4 (Fig. S4g). In both therapies, responsiveness to ICI significantly extended survival.

We then characterized which cell types were predominantly involved in the B16mOVA^HI^ tumor rejection by depleting specific immune cells populations upon ICI treatment. In the absence of the CD4^+^ T cells, CD8^+^ T cells were still capable of controlling tumor growth and led to the rejection in 3 out of 5 mice, thus with slightly lower efficacy than with proper help from the CD4^+^ T cells, as highlighted by the isotype control group in which all tumors were rejected (Fig. 1f). In contrast, CD4^+^ T cells alone were insufficient to eradicate tumors, although they slightly slowed tumor growth as compared to tumors depleted of both CD8^+^ and CD4^+^ T cells. Similarly, we found that NK1.1^+^ cells were not required for responsiveness to ICI (Fig. 1g). Lastly, we found that muMT^-^ transgenic mice, which lack mature B cells and cannot produce IgG, were able to reject B16mOVA^HI^ tumors upon ICI, importantly highlighting that IgG-based antibody-dependent cytotoxicity mechanisms were not necessary for tumor eradication, although we do not exclude that they might take place in WT mice (Fig. 1h).

Finally, we investigated whether the immune rejection of the B16mOVA tumors upon ICI was solely directed against membrane-bound OVA or if immune reactions against other tumor-associated antigens were at play. Upon re-challenge, mice that rejected B16mOVA tumors showed delayed growth of B16 WT tumors, suggesting the presence of pre-existing immune reactions against B16 WT neoantigens induced during the initial rejection of B16mOVA (Fig. 1i, Fig. 4h). Of note, the secondary B16 WT tumors remained non-responsive to ICI. Therefore, while mOVA was necessary to eradicate the primary tumor upon ICI, its loss in the secondary tumors still resulted in delayed tumor growth, potentially mimicking a situation of cancer relapse or metastasis.

### mTMB increase patient survival upon ICI

The remarkable ability of a membrane-bound antigen (i.e., mOVA) to restore responsiveness to ICI in the murine melanoma model encouraged us to validate this hypothesis in cancer patients. Therefore, we analyzed publicly available tumor mutation sequencing data of patients treated or not with ICI, from 3 independent studies by Samstein *et al*.^7^, Hellman *et al*.^8^ and Hugo *et al*.^9^. For each tumor mutated gene detected in patients, we extracted the subcellular localization of its encoded protein from the UniProtKB/Swiss-Prot database^20^ (Supplementary Data 1). We then quantified per patient the number of mutated genes that encode for membrane, cytoplasmic, nuclear, or secreted proteins. Genes that encode proteins expressed at several localizations were classified in all locations, in a non-exclusive manner. We lastly normalized the number of protein-encoding mutated genes at a specific subcellular location to the total number of mutated genes, therefore obtaining proportions of protein-encoding mutated genes per subcellular location (Fig. 2a). These proportions are here called mTMB, cTMB, nTMB or sTMB for membrane, cytoplasmic, nuclear, or secreted -localized TMB, respectively.

**Figure 2.**
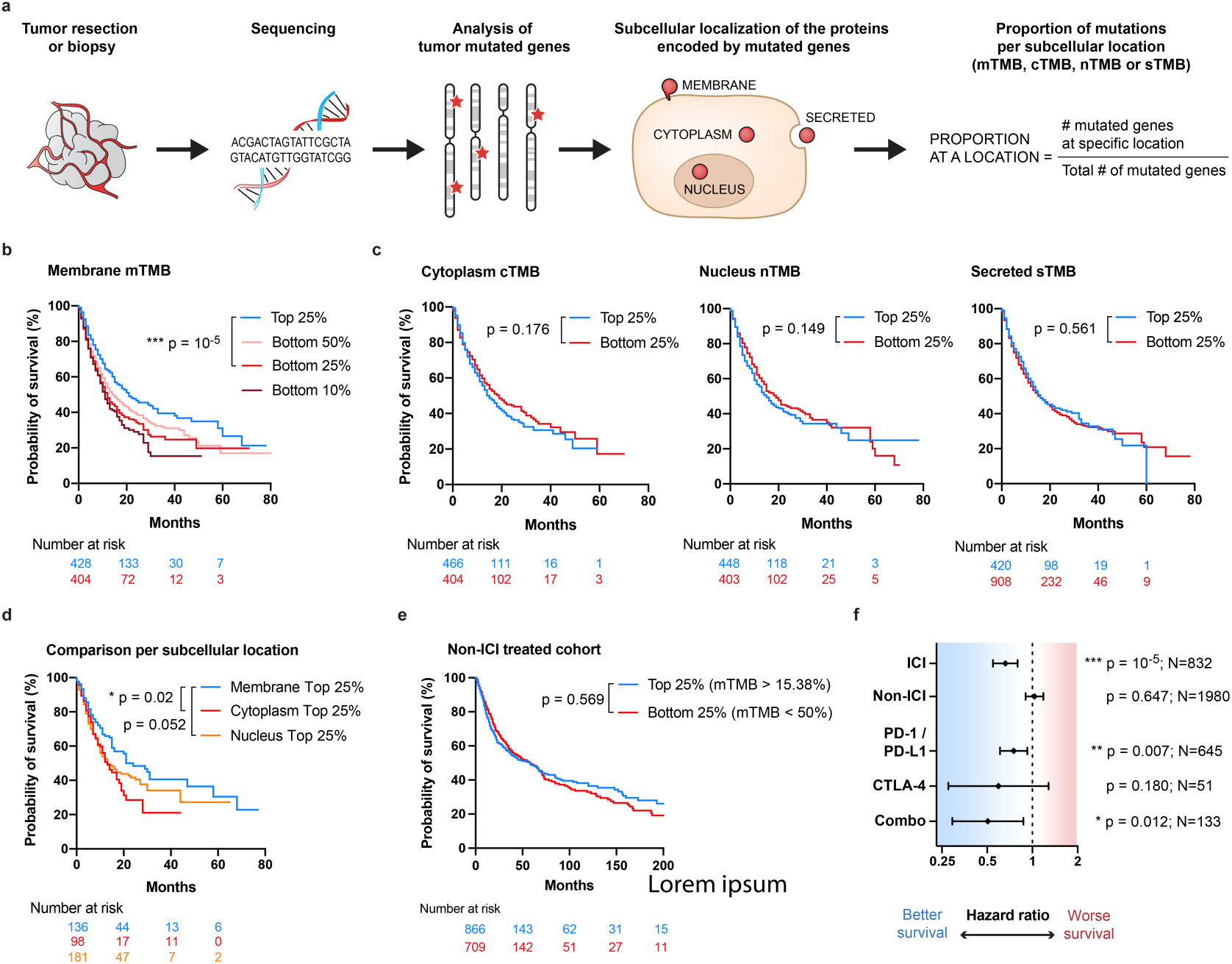
mTMB correlates with an increased survival in cancer patients treated with ICI in a pan-cancer analysis. Data available from Samstein *et al*.^7^. Patients suffering from 9 different cancer types were treated with immune checkpoint inhibitor (ICI) immunotherapy, and their survival was evaluated from the first day of treatment (N=1609 patients). A control cohort of patients non-treated with immunotherapy was used for comparison (N=3142 patients). All Kaplan-Meier survival curves and Cox hazard ratios (HR) for survival were statistically compared using log-rank tests. **a**, Graphical representation of the workflow for the analysis of subcellular localizations associated with the tumor mutations. **b**, Survival of patients with high (Top 25% group) or low (Bottom 50%, 25% or 10% groups) mTMB. **c**, Survival curves of patients having high (Top 25% group) or low (Bottom 25% group) cTMB, nTMB or sTMB. **d**, Survival of patients as a function of their predominant subcellular location of mutated genes (Top 25% groups of membrane, nucleus or cytoplasm mutations; p-values adjusted using Holm-Bonferroni correction). **e**, Survival of non-ICI-treated patients that have high (Top 25%) or low (Bottom 25%) mTMB. **f**, HR for survival of patients having high (Top 25%) versus low (Bottom 25%) mTMB upon ICI treatment, non-treated with immunotherapy (Non-ICI), or depending on the type of ICI received, i.e. PD-1/PDL-1, CTLA-4 or in combination (HR ± 95% CI).

We first analyzed the dataset by Samstein et al.^7^ comprising of 1609 patients with 9 different types of advanced cancers treated with ICI whose tumor mutations were determined using targeted next-generation sequencing MSK-IMPACT (Supplementary Data 2). In total, 424 genes out of the 469 sequenced were classified in the 4 subcellular locations of interest (Fig. S5a). We compared groups of patients with high and low proportion of mutated genes for each specific location using the cutoff values of the upper and bottom group quartiles (Top 25% vs. Bottom 25%; Fig. S5b). A high mTMB was found to correlate with significantly increased patient survival (Fig. 2b). This effect also was conserved at other percentiles than 25% (Fig. S5c). Interestingly, an insufficient mTMB was strongly associated with worsened survival, as highlighted by the gradual decrease between the groups Bottom 50%, 25% and 10%, with the Bottom 10% group being patients with no membrane-localized mutation (Fig. 2b, Fig. S5d). None of the other subcellular locations correlated with significant improvement in survival (Fig. 2c). Instead, trends toward reduced survival were observed for high cTMB and nTMB, and no difference was seen for sTMB. Further division into exclusive patient groups with high proportions of mutated genes at a single location highlighted that the membrane localization provides higher survival benefits than the cytoplasmic and nuclear localizations (Fig. 2d, Fig. S5e).We then questioned whether the survival advantage that correlated with high mTMB was present in non-ICI treated patients. We analyzed 3142 patients from the non-ICI treated control cohort of Samstein *et al*.^7,22^ (Supplementary Data 3) and found that no survival benefit was associated with membrane localization in absence of ICI (Fig. 2e, f). However, all types of ICI therapies, namely PD-1/PD-L1, CTLA-4 or the combination PD-1/PD-L1 + CTLA-4, correlated with extended survival in patients harboring a high mTMB, as indicated by a hazard ratio (HR) for survival inferior to 1. This effect did not reach statistical significance for CTLA-4, likely due to the limited number of patients in this group (Fig. 2f).

Together, these findings suggest that high mTMB improve cancer patient survival upon different types of ICI treatments.

### Impact of mTMB in different cancer types

The ICI-treated cohort analyzed above included patients with 9 different types of cancers, non-equally distributed (Fig. S6a). When comparing the distribution of cancer types within the Top 25% and Bottom 25% of mTMB groups, we noticed that the population with high mTMB was enriched in melanoma, renal cell carcinoma and colorectal cancer patients, and depleted from bladder cancer, glioma and head-and-neck cancer patients (Fig. S6b). This implied that not all cancer types had the same distribution of mTMB; in fact, glioma, bladder and head-and-neck cancers had significantly less mTMB than the pan-cancer group, whereas colorectal and melanoma cancers had significantly more (Fig. 3a).

**Figure 3.**
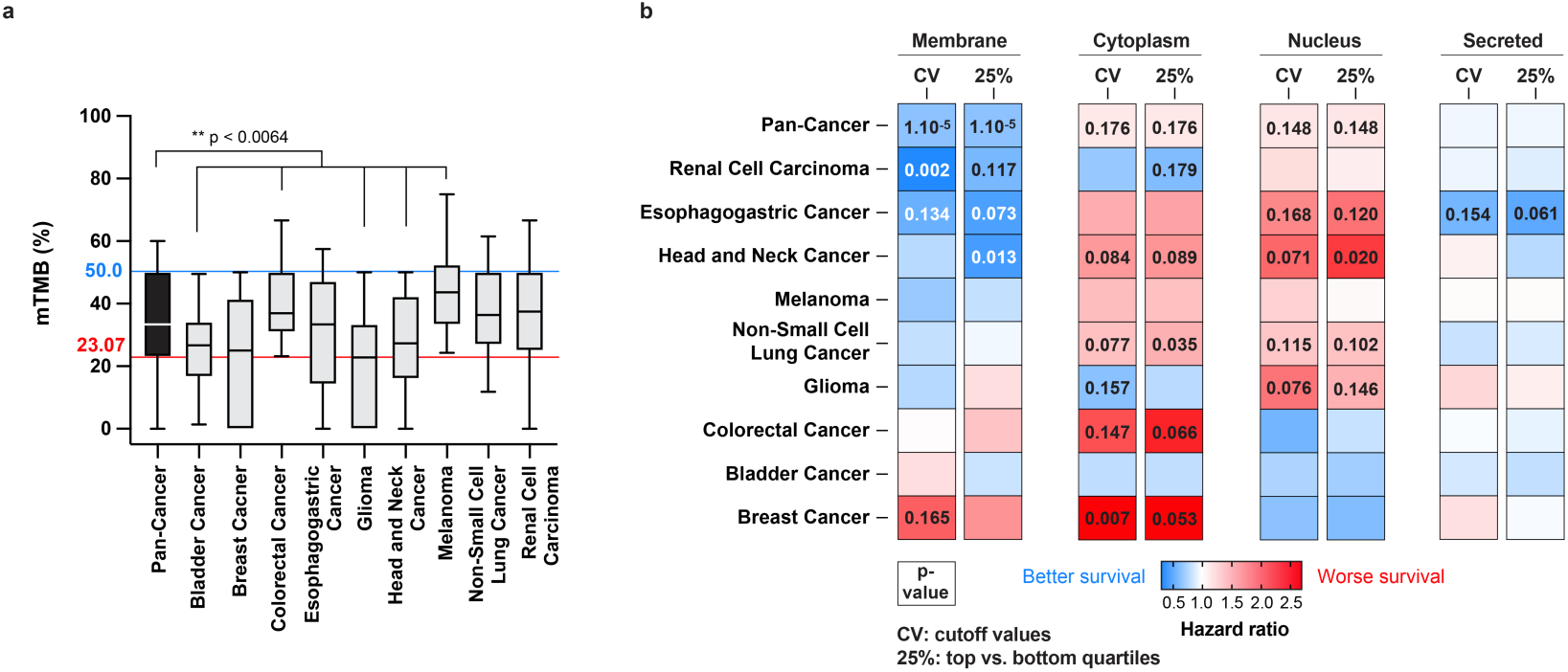
mTMB correlates with increased survival in multiple cancer types. **a**, Distribution of mTMB by cancer types (blue line: cutoff value (CV) for the pan-cancer upper quartile, red line: cutoff value for the pan-cancer lower quartile; Kruskal-Wallis test with Dunn’s post-tests for comparisons to the pan-cancer group). **b**, Heatmap of the HR for survival of patients harboring high versus low mutational load per subcellular location and per cancer types. High and low groups are determined using either the cutoff values (CV) from the pan-cancer group or the upper and lower quartiles (25%) specific to each cancer type (log-rank tests).

Therefore, we detailed the effects of high mTMB, as well as of other subcellular localizations, per cancer type. We computed the HR for survival to compare patients with high versus low proportions of mutated genes at a specific location, using 2 different strategies: 1) keeping the same cutoff values that we used for the pan-cancer group analysis in Fig. 2, reasoning that a “universal” threshold might be determined across cancers as being an absolute proportion of mutations required for extended survival, or 2) using the upper and lower quartile values specific to each cancer type (Fig. 3b, Fig. S6c). Overall, a high mTMB correlates with better survival in 6 out of 9 individual cancers, with statistical significance reached in the renal cell carcinoma and head-and-neck cancer, and close to significance for esophagogastric cancer. The lack of significance in the other cancer types might be due to smaller effects or limited numbers of patients in each sub-cohort. On the other hand, high cTMB and nTMB were associated with worsened survival in a majority of cancer types (6 out of 9; 1 or 2 significantly). Besides, high sTMB did not strongly impact patient survival, except in the esophagogastric cancer, in which a trend toward improvement was observed. Interestingly, both thresholding methods for the selection of high vs. low groups showed very similar results, except for glioma and bladder cancers at the membrane locations. Further analysis with a higher number of patients would clarify whether an absolute threshold for mTMB can be determined to predict increased survival upon ICI across cancers.

### mTMB predict patient response to ICI

While the metric of survival is a relevant measure to evaluate effectiveness of ICI, response rate and long-term survival do not always correlate well. Hence, we searched for published datasets in which the patient response to ICI was reported. We found 2 such studies, from Hellman *et al*.^8^ and Hugo *et al*.^9^, which respectively focused on ICI-treated patients with NSCLC (75 patients) and metastatic melanoma (38 patients). Both studies used whole-exome sequencing (WES) to determine tumor mutations in patients treated with anti-PD-1 or with the combination of PD-1 and CTLA-4 blockade. We thus repeated the subcellular localization analysis using the same algorithm to categories tumor mutated genes according to their possible expression in the membrane, cytoplasm, nucleus or secreted category (Fig. S7a, Supplementary Data 4, 5). Because more genes were sequenced by WES than by MSK-IMPACT, the detected variation range of mTMB in the WES-sequenced patients was much smaller, with most patients having between 25-35% of mutated genes at the membrane location. Interestingly, the overall median mTMB remained similar between the studies, with 33.3%, 27.0% and 34.3% in Samstein *et al*.^7^, Hellman *et al*.^8^ and Hugo *et al*.^9^, respectively (Fig. S7b). The small difference of lowered mTMB found in the cohort from Hellman *et al*.^8^ might be due to the increased number of genes for which the subcellular locations could not be determined.

In this NSCLC cohort^8^, patients that responded to ICI had a significantly higher mTMB, but none of the other studied locations. In addition, these patients tended to survive longer, although statistical significance was not obtained, highlighting the potential discrepancy between ICI responsiveness and overall survival readouts (Fig. 4a, Fig. S7c, d). Impressively, the response rate was 61% in the group with high mTMB (25% Top), vs. 23.5% in patients with low mTMB (25% Bottom) (Fig. 4b). Similar trends were also observed in the melanoma cohort, despite the low number of patients (Fig. S7e-g). Thus, these two additional studies further support the hypothesis that a high mTMB correlates with ICI responsiveness, consistently with the survival results obtained in the larger, multi-cancer cohort from Samstein *et al*.^7^. Importantly, they also point out that this effect was conserved independently of the sequencing methods used for the detection of the tumor mutations.

**Figure 4.**
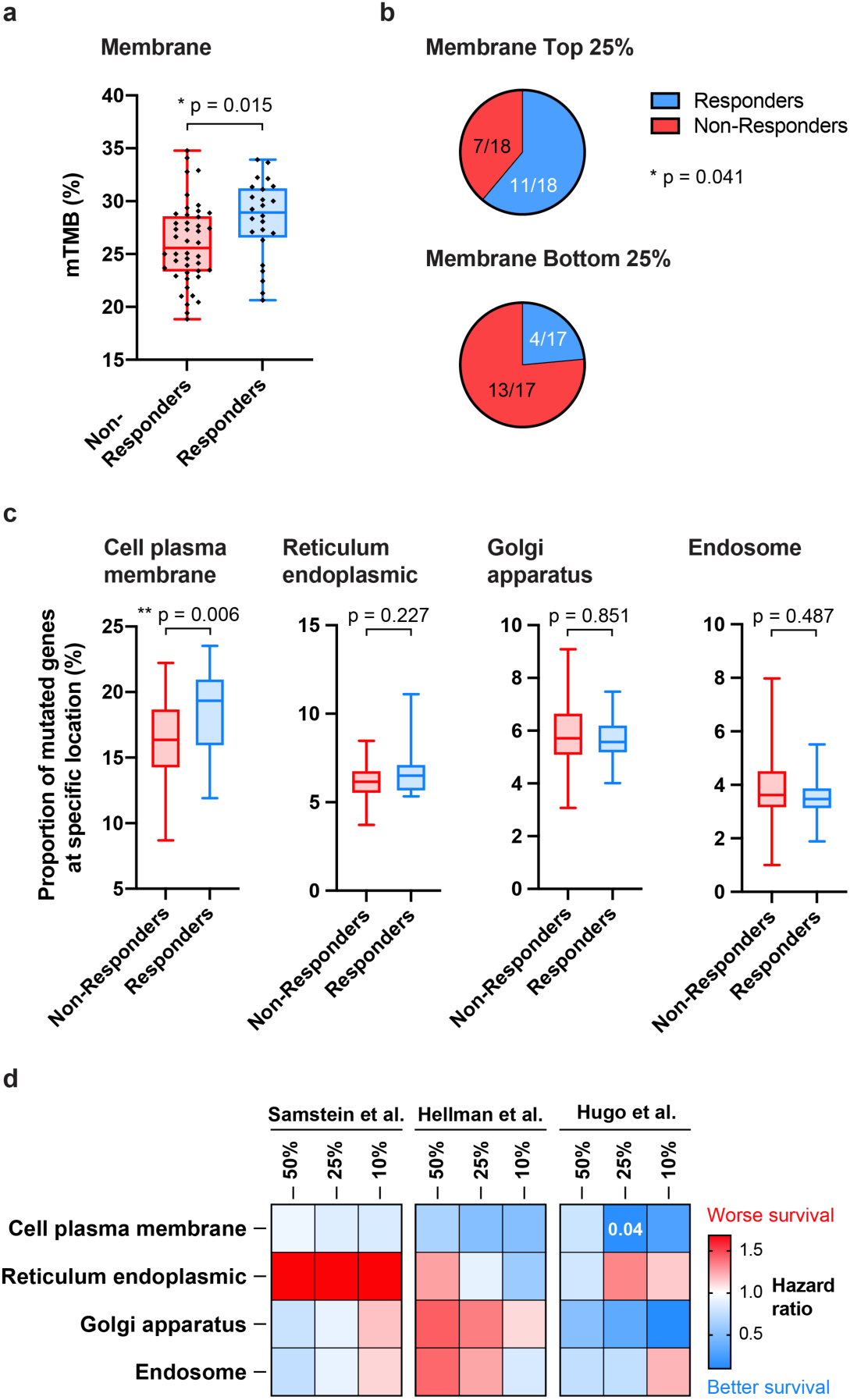
mTMB correlates with better responsiveness to cancer ICI. Data available from Hellman *et al*.^8^. Patients (N=75) with non-small cell lung cancer were treated with a combination of anti-PD-1 and anti-CTLA-4 immunotherapy, and their responsiveness to treatment was evaluated (responders: complete or partial response (CR/PR); non-responders: stable disease or progressive disease (SD/PD)). **a**, mTMB in patients that responded or not to the immunotherapy (Mann-Whitney test). **b**, Proportion of responders and non-responders in patients with high (Top 25%) or low (Bottom 25%) mTMB (Fisher’s exact test). **c**, Proportion of mutated genes at the cell plasma membrane or in other specific membrane-containing cell organelles in responders and non-responders to immunotherapy. **d**, Heatmap of the HR for survival comparing the Top vs. Bottom 50%, 25% or 10% groups having mutations at the plasma membrane or in other membrane-containing organelles, from the cohorts from Samstein *et al*.^7^ (pan-cancer group), Hellman *et al*.^8^ and Hugo *et al*.^9^.

### mTMB at the plasma membrane

Observing that mTMB lead to greater response to ICI, we questioned whether there were differences between particular membranes in the cell. To address this, we refined our algorithm to segregate for cell membrane (i.e., plasma membrane), endoplasmic reticulum, Golgi apparatus or endosome localizations. Using the data on ICI responders from the NSCLC cohort^8^, we found that only the proportion of mutated genes expressing proteins at the cell plasma membrane was significantly increased in ICI responders, while localization at the membranes of organelles did not correlate with changes in ICI response (Fig. 4c). Similar trends were observed for the melanoma cohort (Fig. S7h). In addition, consistent trends toward improvement of survival for patient with increased cell plasma-localized mTMB was observed across the pan-cancer, NSCLC and melanoma cohorts (Fig. 4d, Fig. S7i).

### Membrane-localized mutations as clinical biomarkers for ICI

Finally, we analyzed which membrane protein-encoding mutated genes most impact survival upon ICI. Using the dataset from Samstein *et al*.^7^, we computed the HR of survival between patients bearing mutated and wild-type membrane protein-encoding genes, within each cancer type (Fig. 5a, Supplementary Data 6). We observed that most of the mutated genes correlated with improved survival, although a few of them correlated with worsened survival. We particularly highlighted a subset of 1-13 genes per cancer type for which mutations could serve as potent biomarkers to predict extended survival upon ICI, as indicated by low HRs (in blue in Fig. 5a and Supplementary Data 6). Interestingly, we found that patients bearing at least one of these biomarkers survived significantly longer than patients with none, in all the cancer types for which enough patients were available, i.e., bladder cancer, colorectal cancer, NSCLC, melanoma and renal cell (Fig. S8a). This represents a substantial proportion of patients, between 28.4% and 74.1% depending on the cancer type, thus highlighting a strong potential for clinical translation of these membrane-localized biomarker sets.

**Figure 5.**
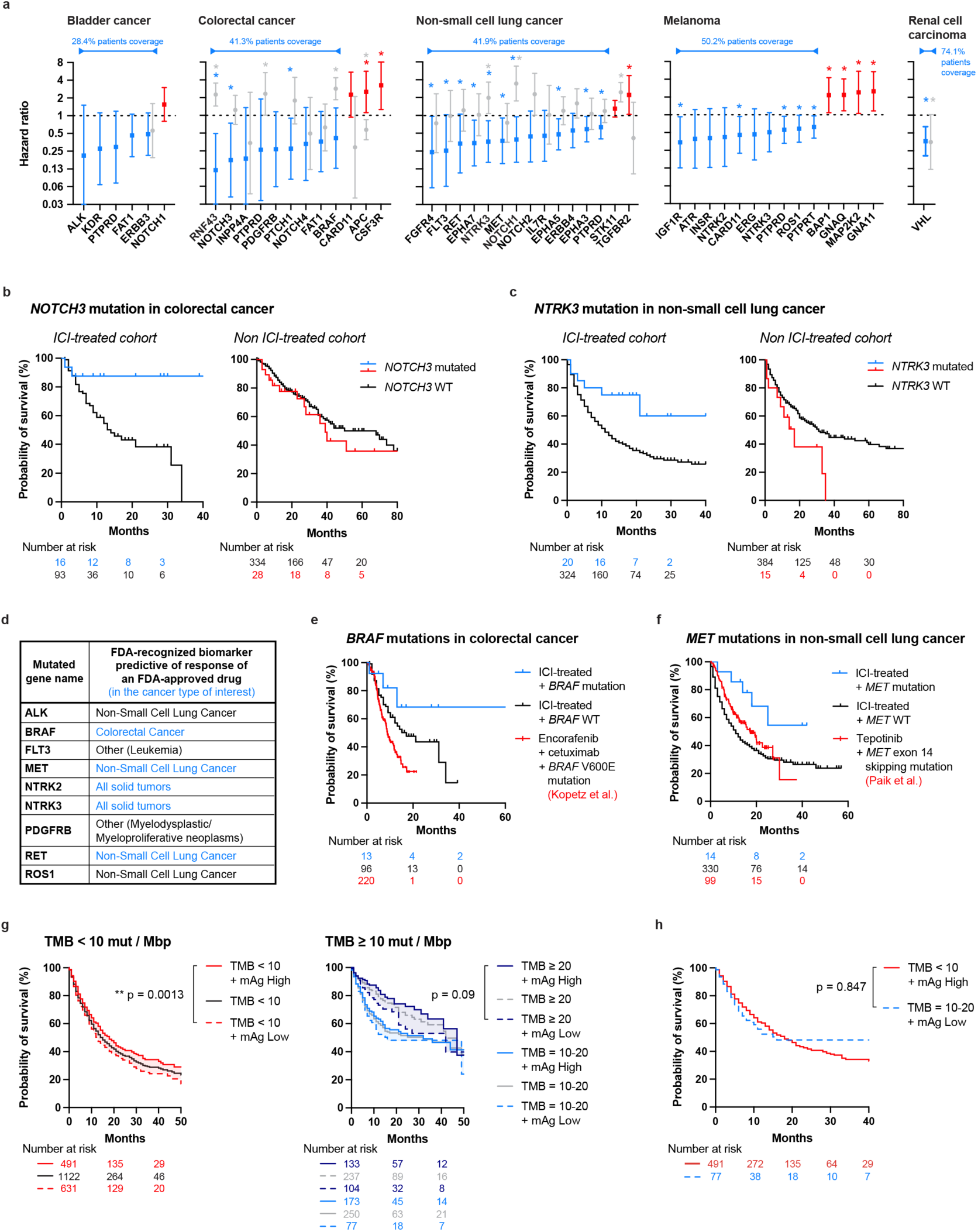
mTMB and specific membrane protein-encoding mutated genes as potent biomarkers for ICI in the clinic. The ICI- and non-ICI-treated cohorts from Samstein *et al*.^7^ were analyzed to determine which membrane protein-encoding mutated genes or combination of them were the most potent to predict survival upon ICI. **a**, HR of survival associated with specific membrane protein-encoding genes per cancer type. A HR < 1 indicates that the mutated version of the gene correlates with increased patient survival as compared to the wild-type gene. The 1-13 membrane-associated top genes for favorable prognosis are labelled in blue, whereas the ones for poor prognosis are in red, for the ICI-treated cohort. Corresponding gene-specific HRs from the non-ICI treated cohort are in grey (log-rank test, *p < 0.05). Patient coverage indicates the proportion of patient that contains at least one of the mutated genes in blue. **b**, Survival curves of ICI and non-ICI treated patients bearing *NOTCH3* mutations in colorectal cancer. **c**, Survival curves of ICI and non-ICI treated patients bearing *NTRK3* mutations in NSCLC. **d**, List of selected membrane protein-encoding genes that are currently recognized by the FDA as biomarkers predictive of a response to FDA approved drug according to the OncoKB database^24^ (those recognized for use within the same cancer type as found in Fig. 5a are shown in blue). **e**, Comparison of survival of patients carrying *BRAF* mutations in colorectal cancer treated with ICI or with the FDA-approved encorafenib+cetuximab therapy (data from Kopetz *et al*.^25^). **f**, Comparison of survival of patients carrying *MET* mutations in NSCLC treated with ICI or with the FDA-approved tepotinib therapy (data from Paik *et al*.^26^). **g**, Comparison of survival of patients with high (> median) or low (< median) mTMB for different levels of TMB (TMB ≥ 10 mut/Mbp being the FDA-validated cutoff for ICI treatment for solid tumors^28^) in a pan-cancer analysis. **h**, Survival curves comparing patients with low TMB (< 10 mut/Mbp) and high mTMB to patients with high TMB (between 10-20 mut/Mbp) and low mTMB, in a pan-cancer analysis. No statistically significant difference was observed between the two groups (log-rank test).

Further seeking ICI-specific membrane-localized biomarkers, we compared the HRs obtained upon ICI to the ones from the non-ICI-treated cohort, for each gene for which enough patients were available (Fig. 5a, Supplementary Data 6). In most cases, gene mutations did not seem to improve survival in the non-ICI-treated cohort to the same extent than in the ICI-treated cohort, suggesting that these biomarkers could be specific for prediction of ICI efficacy. One exception was *VHL* in renal cell carcinoma, for which mutations appeared to be beneficial in both cohorts. On the other hand, we found that some mutated genes correlated with very high survival in the ICI-treated cohort, but with worsened survival in the non-ICI-treated one, such as *NOTCH3* or *RNF43* in colorectal cancer (the latter having been recently elucidated by Zhang et al.^23^), and *NTRK3* and *NOTCH1* in NSCLC (Fig. 5b, c, Fig. S8b, c). Upon confirmation by future studies, such genes could constitute very promising stand-alone biomarkers to guide medical choice toward ICI rather than other treatments in specific cancer types.

Among the membrane-localized biomarkers that we highlighted, a few are recognized by the U.S. Food and Drug Administration (FDA) as biomarkers predictive of response of a FDA-approved drugs^24^ (Fig. 5d), and thus are currently assayed in the clinic. It is the case for some *BRAF* mutations in colorectal cancer, *MET* and *RET* mutations in NSCLC, or *NTRK2/3* mutations^24^. Therefore, we attempted to compare the performance of these membrane-localized biomarkers to predict survival upon ICI versus upon their clinically-associated treatments, using survival data from other published clinical studies by Kopetz *et al*.^25^, Paik *et al*.^26^ and Gautschi *et al*.^27^. While such direct comparisons cannot be conclusive due to intrinsic differences in the study designs, *BRAF* and *RET* mutations seem highly effective as biomarkers for ICI as compared to the FDA-approved encoferanib+cetuximab in colorectal cancer^25^ (Fig. 5e) and tepotinib in NSCLC^26^ (Fig. 5f), respectively. A similar observation was made for *RET* mutations, when compared to a standard-of-care treatment with cabozantinib in NSCLC^27^ (Fig. S8d).

Last but not least, the FDA has very recently approved the use of high TMB (i.e., TMB ≥ 10 mutations/megabase pair (mut/Mbp)) as a criterion for ICI, for adults and children with unresectable or metastatic solid tumors that failed to respond to prior therapies^28^, thus fostering the use of next-generation sequencing of tumor mutations in the clinic. Because determination of the mTMB from these sequencing data would require only a simple algorithm but no additional clinical or laboratory procedures, we examined the benefit of combining mTMB analysis with the standard total TMB analysis to predict survival upon ICI. We found that a high mTMB correlated with improved survival in patients with both low TMB (< 10 mut/Mbp) or high TMB (≥10 mut/Mbp) (Fig. 5g, Fig. S8e, f). In addition, we observed that some patients with high TMB (10-20 mut/Mbp) but low mTMB, for which ICI is approved, had similar survival as patients with low TMB but high mTMB (Fig. 5h), which may not currently qualify for ICI. Importantly, the latter represent 30.5% of the patients in the Samstein *et al*. dataset, which could thus be considered for ICI but would not be otherwise. Together, this suggests that the mTMB could be a valuable parameter to take into account, on top of current TMB analysis, to extend the inclusion criteria for ICI in the clinic.

## DISCUSSION

This study focused on the influence of the subcellular localization of tumor mutations for responsiveness to cancer immunotherapy. Importantly, we demonstrated in both the B16-F10 melanoma mouse model and on a large clinical dataset of 4864 ICI- and non-ICI-treated cancer patients that responsiveness to ICI and extended survival correlated with a high mTMB, especially of mutations localized at the plasma membrane. Interestingly, this effect was not seen for increased load of cytoplasmic, nuclear, or secreted mutations, nor was it seen in patients that were not treated with ICI. This conclusion was supported in a pan-cancer analysis, gathering 9 different types of cancer. While pan-cancer analysis bears the limitation of merging possibly heterogeneous cancer types, it presents the strong advantage of including a large number of patients, therefore increasing statistical power, and mirrors the design of basket clinical trials currently emerging in oncology^29,30^. Further analyses per individual cancer type similarly correlated high mTMB with extended survival in renal cell carcinoma and head and neck cancer in the cohorts from Samstein et al.^7^, although statistical significance depended on the thresholding methods, in a melanoma cohort from Hugo et al.^9^, and in an NSCLC cohort from Hellmann et al.^8^. Nevertheless, in-depth analysis per cancer type would be needed on larger number of patients to further elaborate on these conclusions, as we pointed out that high mTMB might have varying effects in different cancer types.

In our analysis, we found consistent results from clinical datasets published by three independent research groups, which used two different methods of tumor mutation sequencing, namely MSK-IMPACT and WES, both recently approved by the FDA and rapidly emerging in the clinic^31-34^. While these sequencing methods aim to quantify TMB, high load of which is approved as an inclusion criterion for treatment with ICI, our work provides a complementary simple algorithm-based method that can further filter the sequencing data to improve the prediction accuracy of ICI responsiveness. We found that our mTMB criterion indicates 30.5% more patients for inclusion into ICI than the current FDA standard of TMB, based on the Samstein et al. dataset^7^. In addition, we highlighted particular membrane proteins-encoding mutated genes that may be very potent stand-alone predictive biomarkers to guide the choice toward treatment by ICI in certain cancer types.

Although not formally demonstrated here, the membrane-localized proteins encoded by mutated genes constituting the mTMB are likely corresponding to membrane-localized tumor neoantigens. Additional analyses of expression of the mutated genes and prediction of mutated epitope binding on patient-specific MHC molecules would be required to support this assertion. That said, both Hugo et al.^9^ and Hellmann et al.^8^ successfully demonstrated that the total TMB, particularly the amount of somatic non-synonymous single-nucleotide variants (nsSNV), strongly correlates with HLA1 neoantigen load. As to the dataset from Samstein et al., HLA subtypes of patients were not found to be publicly available, to our knowledge, to permit such analysis.

The details of the mechanisms by which membrane-localized mutations modulate immune responses against cancer remain to be clarified. Our data in the B16-F10 mouse model suggest that this effect strongly relies on T cells, rather than on NK cells or on IgG-dependent cytotoxic mechanisms. While CD8^+^ T cells were necessary and sometimes sufficient to eradicate the tumors, our data supports that CD4^+^ T cells provided important help to the CD8^+^ T cells, consistent with other reports that stressed the key role MHC-II restricted neoantigens for responsiveness to ICI^35^. In addition, some previous research has highlighted that secreted^36^, membrane-bound^37^ or extravesicular-bound^38^ antigens enhance CD4^+^ T cell responses and strengthen antigen-specific immunity, in cancer or other contexts. Considering that none of our data supported an immunogenic effect mediated by direct extracellular detection of unprocessed membrane-bound antigens, such as via IgG, we did not further focus on the precise effects of mutations’ localization on cytoplasmic, transmembrane or extracellular domain of the mutated membrane proteins in this study. Lastly, apart from being immune targets, mutations of membrane proteins on tumor cells can impact the primary biological functions of the proteins and their downstream signaling, which could have direct effects on tumor biology, growth and aggressiveness.

Besides the basic immunology perspective, this work provides a rationale for therapeutic immunomodulation by neoantigen selection at different subcellular locations. In particular, personalized cancer vaccines currently target neoantigens based on prediction of MHC binding neoepitopes for optimized T cell activation, with little consideration of the subcellular localization of the neoantigen^13^. Adding vaccinal antigen selection criteria for preferential targeting of plasma membrane neoantigens might improve the therapeutic efficacy of such vaccines. Taken together, we believe that the simplicity of considering the neoantigens’ subcellular localizations for increased predictability to ICI response, the use of mTMB and specific membrane neoantigens as biomarkers to guide medical decisions of cancer treatments, as well as the possible impacts on the design of future immunotherapies, will be valuable in the fight against cancer.

## Data Availability

All data produced in the present work are contained in the manuscript and specific requests can be sent to authors for additional information.

## METHODS

### OVA-expressing B16 melanoma cell lines

B16F10 (B16) melanoma cells (American Type Culture Collection, Manassas, VA, USA) were genetically modified by transduction with OVA-encoding lentivirus. Briefly, OVA-encoding DNA sequences were purchased from GenScript (Piscataway, NJ, USA). In one design, full-length OVA (UniprotKB P01012) was fused at the N-terminus to the signal peptide of mouse H-2K^B^ (aa1-aa21, UniprotKB P01901) and at the C-terminus to the transmembrane domain of mouse H-2D^B^ (aa299-aa331, UniProtKB P01899). Sequences were subcloned in the pLV-mCherry backbone (Addgene #36804) in place of mCherry. Lentiviruses were made by polyethylenimine (PEI)-mediated transfection of human embryonic kidney (HEK) 293-T cells using OVA-encoding plasmid with the packaging plasmids pMD2.G (Addgene #12259), pMDLg/pRRE (Addgene #12251) and pRSV-Rev (Addgene #12253). Twelve hours after transfection, the cell culture medium was refreshed and 36 h later, the medium was collected and filtered at 0.22 μm. Lentiviruses were concentrated by ultracentrifugation at 100,000 xg for 2 h at 4°C and resuspended in phosphate-buffered saline (PBS). B16 cells cultured in 48-well plates were transduced by adding OVA-encoding lentiviruses in the culture medium and centrifuging at 1150 xg for 30 min at room temperature, and then were cultured for 24 h, after which the medium was refreshed. For B16mOVA^HI/LO^ and B16-OVA^HI^, monoclonal selection was performed by limiting dilution, and OVA-expression was quantified by quantitative polymerase chain-reaction (qPCR). The B16-OVA^LO^ cell line was a gift from B. Huard (University of Geneva, Switzerland). All cell lines were tested as negative for mycoplasma contamination by PCR.

### Quantitative PCR for OVA expression

Expression of OVA in B16 cell lines or tumors was quantified by qPCR. Prior to RNA extraction, 30-50 mg of tumor tissues were homogenized (FastPrep-24 5G, MP Biomedicals, Santa Ana, CA, USA) in RLT lysis buffer (Qiagen, Hilden, Germany), spun down at 10,000 xg for 10 min and the supernatant was collected. For cells in culture, 1-2 million cells were pelleted, washed with PBS and lysed in RLT buffer. RNA was extracted using the RNeasy Plus Mini kit (Qiagen). The extracted RNA (1 μg) was then converted to cDNA using SuperScript IV VILO Master Mix (ThermoFisher Scientific, Waltham, MA, USA). All kits were used according to manufacturers instructions. TaqMan qPCR were finally performed using TaqMan Universal PCR Master Mix, OVAL primer (Gg03366807_m1) and ActB primer (Mm02619580_g1) (ThermoFisher Scientific), in a LightCycler 96 real-time PCR system (Roche Life Science, Basel, Switzerland).

### Detection of membrane-bound OVA

Surface-expression of OVA was verified by flow cytometry and microscopy. Single cell suspensions of the different OVA-expressing B16 were incubated for 30 min on ice with anti-OVA (ab181688, Abcam, Cambridge, UK) in PBS + 2% fetal bovine serum (FBS). Cells were washed twice and stained using an anti-rabbit secondary antibody (A315723, Invitrogen, Carlsbad, CA, USA) for 20 min on ice in the dark. Cells were washed and analyzed by flow cytometry (BD LSRFortessa, BD Biosciences, Franklin Lakes, NJ, USA) or imaged by fluorescence microscopy (Leica DMi8, Wetzlar, Germany). Flow cytometry data were analyzed using FlowJo (FlowJo LLC) and microscopy images were processed using Fiji (ImageJ, U.S. National Institutes of Health, Bethesda, MD, USA).

### Extracellular vesicles (EV) isolation

EV from the B16mOVA^HI^ and B16-OVA^HI^ cell lines were harvested using the CLAD1000 system (2440655, Cole-Parmer, Vernon Hills, IL, USA) as described by Mitchell *et al*. ^39^. Briefly, 16 million cells were suspended in 15 mL complete EV-depleted DMEM (DMEM + 1% penicillin/streptomycin (P/S) + 10% exosome-depleted FBS (A2720801, Thermo Fisher Scientific)) and loaded into the lower chamber of the CLAD flask. The upper chamber was then loaded with DMEM + 1% P/S, and cells were allowed to recover for 4 days. On the 4th day, the upper reservoir was emptied and the media in the lower chamber was collected. The lower chamber was washed twice with DMEM, collecting only the first wash. The lower chamber was then refilled with 15 mL of complete EV-depleted DMEM. This harvesting process was repeated every 4 days. Collected media was first spun at 300 xg for 10 min to remove cells, then centrifuged at 3000 xg for 10 min to remove large cell debris and finally at 10,000 xg for 30 min to further remove debris. The final supernatant was concentrated using 100,000 MWCO concentrator tubes (UFC910024, EMD Millipore, Burlington, MA, USA) before processing via size exclusion. Size exclusion was performed using the Izon qEV10 system (IZON SP3) according to the manufacturer’s instructions to collect separately the EV fractions, containing particulates of 70-1000 nm in size, and the non-particulates non-EV fractions. Once purified, EV harvests were pooled and re-concentrated. Total protein content of the purified EV was quantified using a Micro BCA kit (Thermo Fisher) before storage at -20°C. Equal amount of proteins (34 μg) were loaded on SDS-PAGE gels for further analysis by western blot.

### Western blot analysis

Samples were run on SDS-PAGE gels for 45 min at 140 V (Mini-PROTEAN gel system, Bio-Rad Laboratories, Hercules, CA, USA) in Laemmli loading buffer before being transferred onto western blot membranes (Immobilon-P PVDF membrane, EMD Millipore; Mini Trans-Blot cell, Bio-Rad) for 1 h at 90 V. Membranes were blocked using 5% milk in PBS + 0.05% Tween-20 (PBST) overnight at 4°C under agitation and probed with anti-OVA (ab181688) for 4 h at room temperature. Membranes were washed in PBST thrice and incubated with a horseradish peroxidase (HRP)-conjugated anti-rabbit secondary antibody for 1 h at room temperature. Membranes were washed at least 3 times for 5 min in PBST, revealed using the Clarity Western ECL substrate (Bio-Rad) and imaged using a gel imaging system (Universal Hood III, Bio-Rad).

### Mice

All animal experimentation was approved by the University of Chicago Institutional Animal Care and Use Committee in compliance with local ethical and procedural regulations. Mice were purchased from The Jackson Laboratory (Bar Harbor, ME, USA). Female C57BL/6J (No 000664) or female MuMt^-^ mice (B6.129S2-*Ighm*^tm1Cgn^/J, No 002288) were between 8-12 weeks old at the start of the experiments, with mice being aged-matched within an experiment. Act-mOVA mice (C57BL/6-Tg(CAG-OVAL)916Jen/J, No 005145) were bred in-house and female mice of 25-35 week old were used for experimentation. Mice were housed at the Animal Resources Center Facility at the University of Chicago, had water and food *ad libitum*, and were daily monitored for health care.

### *In vivo* antibodies

All antibodies used *in vivo* were the InVivoMAb grade antibodies purchased from Bio X Cell (Lebanon, NH, USA). Antibodies used as immune checkpoint therapies were anti-PD-1 (clone 29F.1A12), anti-PD-L1 (clone 10F.9G2) and anti-CTLA-4 (clone 9H10). Antibodies used for immune cell depletion were anti-CD8α (clone 2.43), anti-CD4 (clone GK1.5), anti-NK1.1 (clone PK136), Isotype IgG2a (clone C1.18.4), Isotype IgG2b (clone LTF-2).

### Tumor injections

Mice were anaesthetized by isoflurane inhalation and were injected intradermally with 1.5 million of the different OVA-expressing or WT B16 cell lines. The tumor was measured using a digital caliper every 2 days, and tumor volume was calculated as follows: volume = length*width*height*(π/6). Mice were euthanized if sick or when the tumor volume reached 1 cm^3^. When indicated, mice were treated with immunotherapy, i.e. anti-PD-1 (200 μg) or the combination anti-PDL-1 + anti-CTLA-4 (100 μg each), once by intraperitoneal injection when the tumor volume was between 20-50 mm^3^ (day 5-8 post-tumor injection). When needed, 500 μg of depletion antibodies (anti-CD8α, anti-CD4, anti-NK1.1 or isotype control) were injected intraperitoneally 24 h after the checkpoint inhibitor therapy and repeated 7 days later. In the re-challenge experiments, 250k WT B16 cells were injected intradermally on the contralateral side on the mice 1 month after they cleared the primary tumor.

### Flow cytometry analysis of tumor

Ten days after tumor injection, tumor were harvested on euthanised mice. Tumors were weighed, and about 300 mg were processed. Tumors were cut into small pieces, digested for 45 min in collagenase IV (1 mg/mL), DNAse I (40 μg/mL) in DMEM + 2% FBS + 1.2 mM CaCl_2_ at 37°C under magnetic stirring. The samples were pipetted 100 times to dissociate tumor pieces, and single cell suspensions were obtained by using 70 μm cell strainer. Cells were kept on ice. Undigested pieces were further mixed with collagenase D (3.3 mg/mL), DNAse I (40 μg/mL) in DMEM + 2% FBS + 1.2 mM CaCl_2_ for 30 min at 37°C and collected as above. EDTA (5 mM) was added to the single cell suspension. The equivalent of 20 mg of tumor was used for staining for flow cytometry analysis. Tumor samples were washed in PBS and stained for cell viability for 15 min using Fixable Viability Dye eFluor 455UV (eBioscience, San Diego, CA, USA). The cells were washed and Fc receptors were blocked using anti-CD16/32 (#101302, BioLegend) for 20 min. Cells were then stained for 20 min on ice using the following antibodies: anti-CD45 (30-F11), anti-CD8α (53-6.7), anti-PD-1 (29F.1A12), anti-NK1.1 (PK136), anti-Ly6G (1A8), anti-Ly6C (HK1.4), anti-CD11b (M1/70), anti-F4/80 (BM8), anti-I-A/I-E (M5/114.15.2), from BioLegend (San Diego, CA, USA); anti-CD3ε (145-2C11), anti-CD4 (GK1.5), anti-CD62L (MEL-14), anti-CTLA-4 (UC10-4F10-11), anti-CD25 (PC61), anti-CD80 (16-10A1), anti-B220 (RA3-6B2), anti-CD19 (1D3), anti-CD11c (HL3), from BD Biosciences; anti-CD44 (IM7), anti-CD103 (2E7), from eBioscience. Cells were washed before analysis. When needed, intracellular staining with anti-FoxP3 (MF23, BD Biosciences) was performed using the BD Cytofix/Cytoperm Plus kit (BD Biosciences) according to the manufacturer’s instruction. All staining procedures were done on ice with samples protected from light, in PBS + 2% FBS + 1 mM EDTA when not stated otherwise. Cells were analyzed using a LSRFortessa flow cytometer (BD Biosciences). Data were processed using FlowJo (FlowJo LLC). Gating strategies for the flow analysis and biomarkers used to define cell populations are detailed in Supplementary Data 1.

### *Ex vivo* antigen-specific T cell restimulation

Ten days after tumor injection, spleens were harvested on euthanized mice. Single cell suspensions of splenocytes were obtained using a 70 μm cell strainer. Cells were washed in PBS before the red blood cells were lysed in ACK buffer (Lonza, Basel, Switzerland) for 4 min and blocked with complete media (IMDM + 10% FBS + 1% P/S). Cells were centrifuged, resuspended in complete media, and 0.5 million were plated in in 96 U-bottom plate. OVA_257-264_ (SIINFEKL; GenScript) and OVA_323-339_ (ISQAVHAAHAEINEAGR; GenScript) were added to the splenocytes at a final concentration of 1 μg/mL to restimulate CD8^+^ and CD4^+^ T cells, respectively. Unstimulated controls were tested using complete media without peptide, and positive controls were tested using ionomycin (1 μg/mL) + PMA (50 ng/mL). After 4 days in culture, the cell supernatant was collected and the amount of IFNγ secreted was quantified using mouse IFNγ quantikine ELISA kit (R&D systems, Minneapolis, MN, USA) according to the manufacturer’s instructions. Data represent the concentration of IFNγ secreted in restimulated culture supernatants subtracted with the amount detected in unstimulated supernatants.

### IgG titration in plasma

Ten days after tumor injection, mice were bled by intracardiac puncture upon euthanasia. The blood was collected in EDTA-containing tubes, spun down at 1000 xg for 5 min and the plasma was collected and stored at -80°C until analysis. ELISA plates (Maxisorp, Nunc, Roskilde, Denmark) were coated with 10 μg/mL OVA (Sigma-Aldrich, St. Louis, MO, USA) in PBS overnight at 4°C, and blocked with casein (Sigma-Aldrich, St. Louis, MO, USA) for 2 h at room temperature. The plates were washed with PBST, and plasma diluted in casein was added to the wells, starting at a concentration of 1:100 and serially diluted by 10, for 2 h at room temperature. The plates were washed again, and the following HRP-conjugated antibodies were used for detection: anti-mouse IgG1 (#1070-05), anti-mouse IgG2a (#1080-05), anti-mouse IgG2b (#1090-05) and anti-mouse IgG3 (#1100-05) from Southern Biotech (Birmingham, AL, USA). The plates were revealed with TMB substrate (EMD Millipore) and stopped with 2N H_2_SO4. Absorbance at 450 nm was read using an Epoch ELISA reader (BioTek, Winooski, VT, USA), and corrected by the absorbance at 570 nm. Antibody titers were determined as the highest plasma dilution for which the corrected absorbance was twice the background level. The area under curve (AUC) was calculated as area under the titration curve of the log_10_(corrected absorbance over background).

### Human data analysis

Processed sequencing data of tumor mutations (list of tumor mutated genes and tumor mutational burden score) and corresponding patient clinical data were obtained from the studies by Samstein *et al*.^7^, Hellman *et al*.^8^, and Hugo *et al*.^9^. Subcellular locations associated with *Homo Sapiens* genes (taxon ID = 9606) were uploaded from UniProtKB/Swiss-Prot database on August 2, 2020 and are provided in Supplementary Data 2 (Gene subcellular locations inventory). Algorithms for data processing and analysis were coded in R (Rstudio, Boston, MA, USA).

For each distinct tumor mutated gene of a patient, we searched the gene name that matches in the gene subcellular locations inventory file. Genes that were not found were categorized as “Unfound genes”. Genes that were found but for which the subcellular location was unfound were further checked on the online UniprotKB/Swiss-Prot database using the GetSubcellular_location() function from the R package ‘UniprotR’. If the gene subcellular location remained unfound, it was then categorized as “Unknown location”. When multiple locations were found for a specific gene name, they were concatenated to obtain a single subcellular location entry per gene name. The gene subcellular locations were then categorized as membrane, cytoplasmic, nuclear or secreted by checking the presence of the character sequence: membrane = “Membrane” or “Cell membrane”, cytoplasmic = “Cytoplasm”, nuclear = “Nucleus”, secreted = “Secreted” in the subcellular location entry associated with the gene. When indicated, the categories were extended to the cell membrane = “Cell membrane”, the endoplasmic reticulum = “Reticulum” or “reticulum”, the Golgi apparatus = “Golgi” or “golgi”, or endosomal location = “Endosome” or “endosome” or “Endosomal” or “endosomal”. Oftentimes, a single gene name was associated with several subcellular locations, in which case the gene was included in several category in an non-exclusive way. For each patient, we counted the number of mutated genes at each specific subcellular locations, and the proportion of mutated genes at a specific location was computed as the “number of tumor mutated genes at a location divided by the total number of tumor mutated genes in the patient”. Patients with no tumor mutated genes were removed from the analysis. In the presented data, groups of patients were determined using inclusive percentiles, except in groups separated at the median, for which the group below median was inclusive and the group above median was exclusive. In Fig. 5, “mTMB High” and “mTMB Low” correspond to groups of patients for which mTMB was respectively higher and lower than the median.

In R, survival analysis were performed using the libraries ‘survival’, ‘survminer’ and ‘survcomp’ and the functions survfit() and surv_pvalue(). Hazard ratios were computed using the function hazard.ratio(). All these functions were used using log-rank tests when asked. In case of multiple comparisons, p-values were adjusted using the function p.adjust().

### Analysis of the membrane-localized biomarkers

HR of survival was computed for each membrane protein-encoding gene, between patients that bear mutated version of the gene vs. patients that bear the wild-type version of the gene. The analysis was done independently for each cancer type. Results were considered relevant to report (in Fig. 5) when at least 7 patients with a mutated version of a gene were available, and 1) when the HR was ≤ 0.5 or statistically significance by the log-rank test was reached, or 2) when the HR ratio was ≥ 1.3 and close to statistically significance (p-value < 0.2). The non ICI-treated cohort results were reported when at least 7 patients had the mutated version of the gene of interest.

### Statistics & Software

Graphs were plotted using Prism 9 (GraphPad, San Diego, CA, USA). Statistical analysis were run on Prism 9 or on R (RStudio). Overall threshold for statistical significance was considered as p-value < 0.05. Figures were made on Illustrator CS5 (Adobe, San Jose, CA, USA).

### Material, data and code availability

Tumor mutation sequencing data for the human cohorts used in this study are publicly available from Samstein *et al*.^7^, Hellman *et al*.^8^ and Hugo *et al*.^9^. Subcellular locations associated to *Homo Sapiens* genes are provided in Supplementary Data 1 and updated versions can be downloaded from the UniProtKB/Swiss-Prot database. Proportion of mutation subcellular locations per patient and corresponding selected clinical data are provided in Supplementary Data 2-5. Other material, data and code remains available upon request to the corresponding authors.

## SUPPLEMENTARY INFORMATION

***Supplementary Data 1:*** Subcellular locations of proteins associated with *Homo Sapiens* genes.

***Supplementary Data 2:*** Proportion of mutations at specific location for ICI-treated cohort from Samstein *et al*.^7^

***Supplementary Data 3:*** Proportion of mutations at specific location for the non-ICI-treated cohort from Samstein *et al*.^7^

***Supplementary Data 4:*** Proportion of mutations at specific location for the NSCLC patients cohort from Hellman *et al*.^8^

***Supplementary Data 5:*** Proportion of mutations at specific location for the melanoma patients cohort from Hugo *et al*.^9^

***Supplementary Data 6:*** HR of survival per mutated genes and per cancer type for the ICI and non-ICI treated patients cohort from Samstein *et al*.^7^

## ACKNOWLEDGMENTS

The authors would like to thank Prof. Luc G. T. Morris and Prof. David B. Solit for their help in the use of their clinical dataset, Dr. Raga Krishnakumar, Dr. Alexandre de Titta and Dr. Sandra Gribi for advice on data analysis and Dr. Jialu Liu for technical assistance. This article is dedicated to the memory of Anthony Gomes. This work was funded by the Chicago Immunoengineering Innovation Center of the University of Chicago and NIH R01 CA219304 (to M.A.S.).

## AUTHOR CONTRIBUTIONS

P.S.B., S.H., M.A.S. and J.A.H. have conceived the project and designed the experiments. P.S.B., S.H., Z.G., T.K., A.T.A., G.R., Y.W. and S.G., performed the experiments. P.S.B., S.H., Z.G., T.K, P.B and J.A.H. analyzed and interpreted the data. P.S.B, J.A.H., and Z.G. wrote the manuscript, and S.H., P.B. and M.A.S. corrected it.

## COMPETING INTERESTS

The University of Chicago has filed for patent protection for biomarkers described in this work, upon which P.S.B., Z.G., S.H. and J.A.H. are inventors. Other authors have no competing interest to declare.

## Notes

### Author Declarations

The study used only publicly available sequencing data from human patients, published by Samstein et al., Hugo et al, and Hellman et al. as described in the manuscript.

