## Supplementary Data for "Membrane-Localized Mutations Predict the Efficacy of Cancer Immunotherapy"

### SUPPLEMENTARY INFORMATION

***Supplementary Data 1:*** Subcellular locations of proteins associated with *Homo Sapiens* genes.

***Supplementary Data 2:*** Proportion of mutations at specific location for ICI-treated cohort from Samstein *et al.*[7]

***Supplementary Data 3:*** Proportion of mutations at specific location for the non-ICI-treated cohort from Samstein *et al.*[7]

***Supplementary Data 4:*** Proportion of mutations at specific location for the NSCLC patients cohort from Hellman *et al.*[8]

***Supplementary Data 5:*** Proportion of mutations at specific location for the melanoma patients cohort from Hugo *et al.*[9]

***Supplementary Data 6:*** HR of survival per mutated genes and per cancer type for the ICI and non-ICI treated patients cohort from Samstein *et al.*[7]

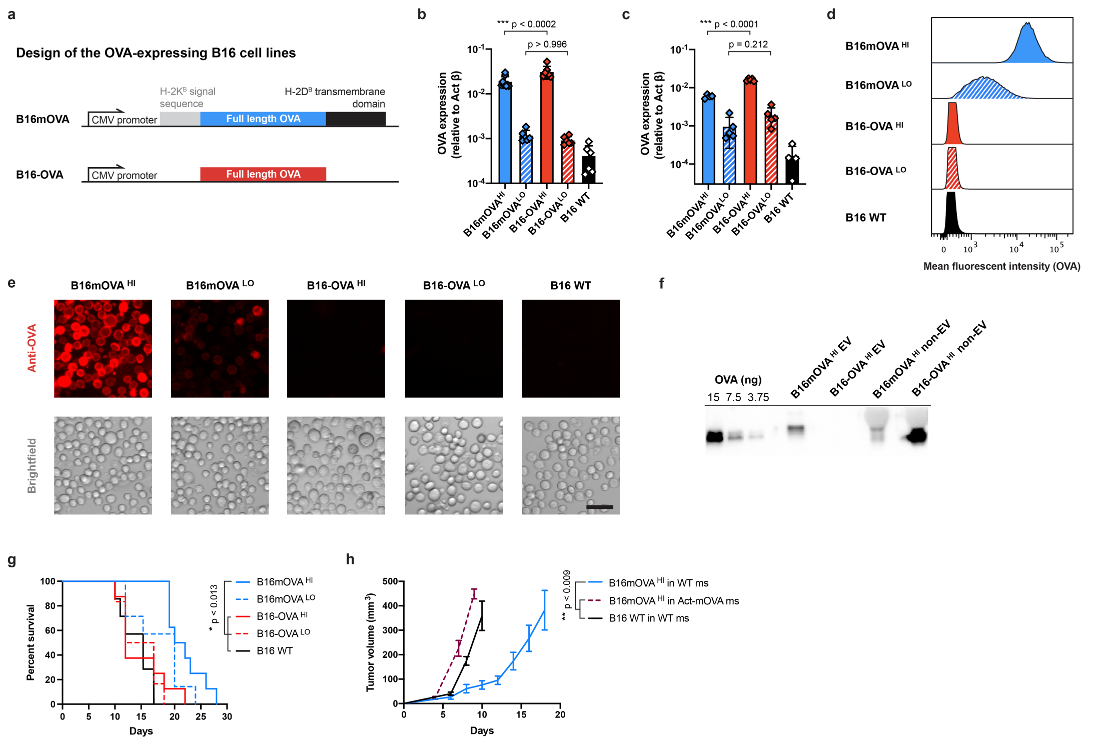


***Supplementary Fig. 1.*** **Characterization of OVA-expressing B16-F10 melanoma cell lines and tumors. a,** Design of the different OVA-expressing B16-F10 cell lines, expressing membrane OVA (mOVA) or soluble OVA. **b,** OVA expression in the modified B16 cell lines in culture *in vitro*, assessed by qPCR (N≥6, mean ± SD, ANOVA with Sidak's post-test). **c,** OVA expression in the modified B16 tumors *in vivo*, assessed by qPCR (N≥4, mean ± SD, ANOVA with Sidak's post-test). **d,** Cell-surface staining of OVA quantified by flow cytometry via the mean fluorescence intensity. **e,** Detection of cell plasma membrane-bound OVA on the different OVA-expressing B16 cell lines assessed by microscopy (red: anti-OVA; scale bar = 50 µm). **f,** Western blot analysis for OVA detection in the extracellular vesicles (EV) produced *in vitro* by B16mOVA$^{HI}$ or B16-OVA$^{HI}$ cells lines or in the non-EV fraction (black = positive detection of OVA). **g,** Survival of mice injected with the different OVA-expressing cell lines, associated to the tumor growth curves of Fig. 1a (N≥8, log-rank tests with Holm-Bonferroni p-values adjustment). **h,** Tumor growth of B16mOVA$^{HI}$ in Act-mOVA mice as compared to growth in wild-type (WT) mice (N≥3, mean ± SEM, Kruskal-Wallis with Dunn's post-test at day 10).

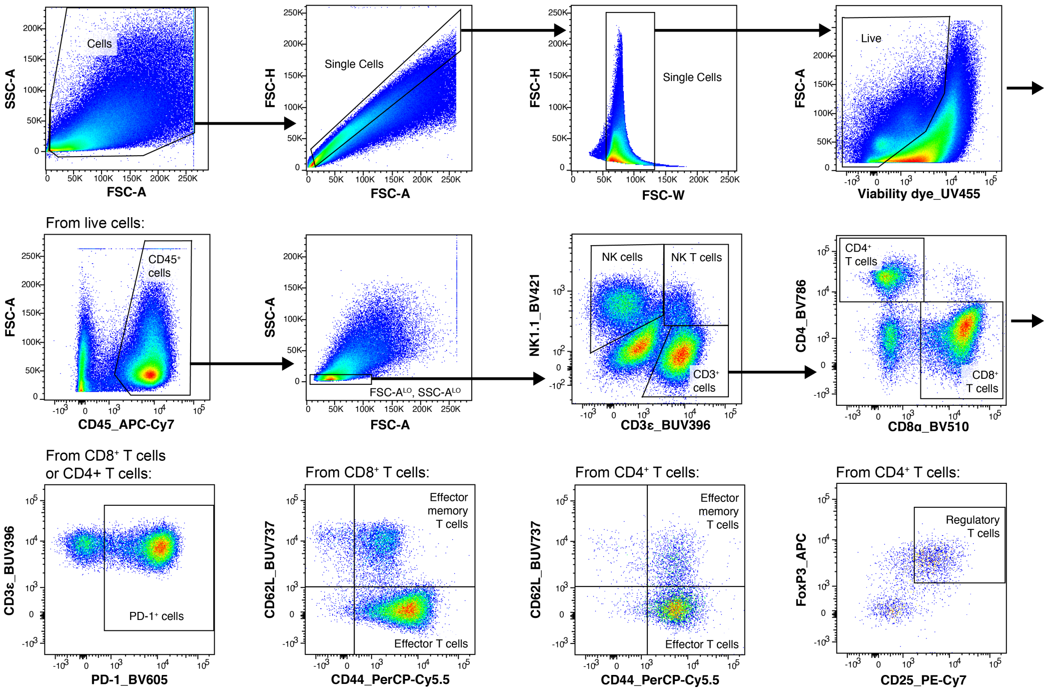


***Supplementary Fig. 2.*** **Gating strategy for the characterization of T and NK cells.** Multi-colored flow cytometry was used to analyze the subsets of T and NK cells in the tumors at day 10 post-injection. Subset of immune cells were defined using the following markers: NK cells (FSC$^{LO}$, SSC$^{LO}$, CD45$^{+}$, NK1.1$^{+}$, CD3ε$^{-}$), NK T cells (FSC$^{LO}$, SSC$^{LO}$, CD45$^{+}$, CD3ε$^{+}$, NK1.1$^{+}$), CD8$^{+}$ T cells (FSC$^{LO}$, SSC$^{LO}$, CD45$^{+}$, NK1.1$^{-}$, CD3ε$^{+}$, CD8$^{+}$), CD4+ T cells (FSC$^{LO}$, SSC$^{LO}$, CD45$^{+}$, NK1.1$^{-}$, CD3ε$^{+}$, CD8$^{+}$), effector T cells (same markers than T cells with CD44$^{+}$, CD62L$^{-}$), effector memory T cells (same markers than T cells with CD44$^{+}$, CD62L$^{+}$). regulatory T cells (same as CD4$^{+}$ T cells with CD25$^{+}$, FoxP3$^{+}$).

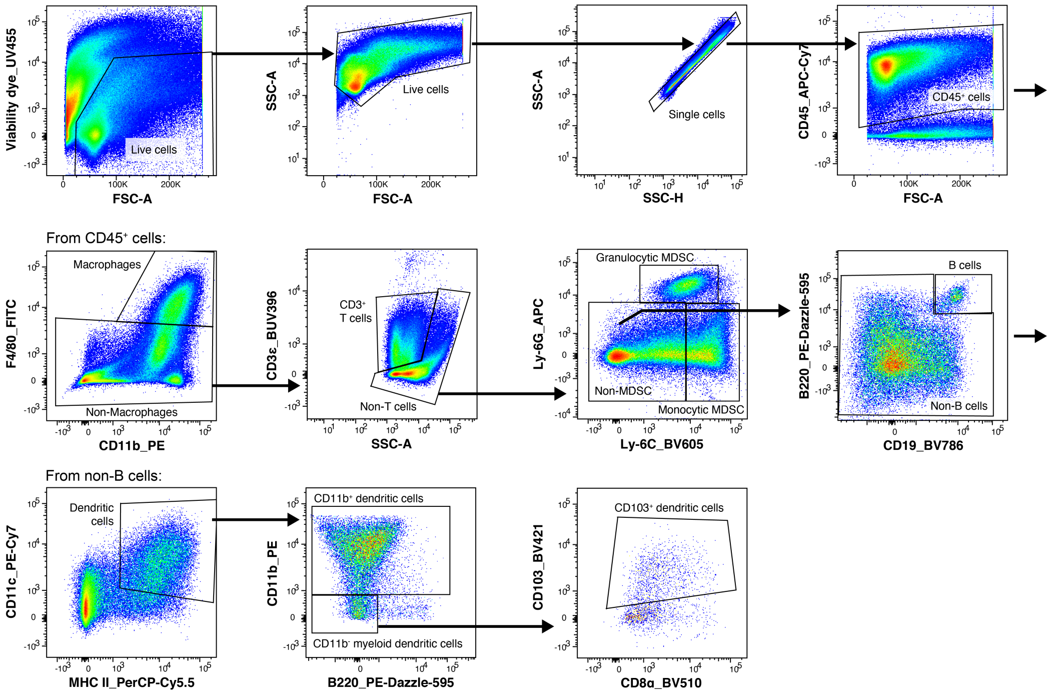


***Supplementary Fig. 3.*** **Gating strategy for the characterization of B cells and myeloid cell subsets.** Multi-colored flow cytometry was used to analyze the subsets of B cells and myeloid cells in the tumors at day 10 post-injection. Subset of immune cells were defined using the following markers: Macrophages (CD45$^+$, F4/80$^+$, CD11b$^+$), Granylocytic myeloid-derived suppressor cells (MDSC) (CD45$^+$, F4/80$^-$, CD3ε$^-$, Ly6G$^+$, Ly6C$^{MID/HI}$), Monocytic MDSC (CD45$^+$, F4/80$^-$, CD3ε$^-$, Ly6G$^-$, Ly6C$^{HI}$), B cells (CD45$^+$, F4/80$^-$, CD3ε$^-$, Ly6G$^-$, Ly6C$^{LO/MID}$, CD19$^+$, B220$^+$), dendritic cells (DCs) (CD45$^+$, F4/80$^-$, CD3ε$^-$, Ly6G$^-$, Ly6C$^{LO/MID}$, CD11c$^+$, MHCII$^+$), CD11b$^+$ DCs (same than DCs with CD11b$^+$), CD103$^+$ DCs (same than DCs with CD11b$^-$, B220$^-$, CD103$^+$).

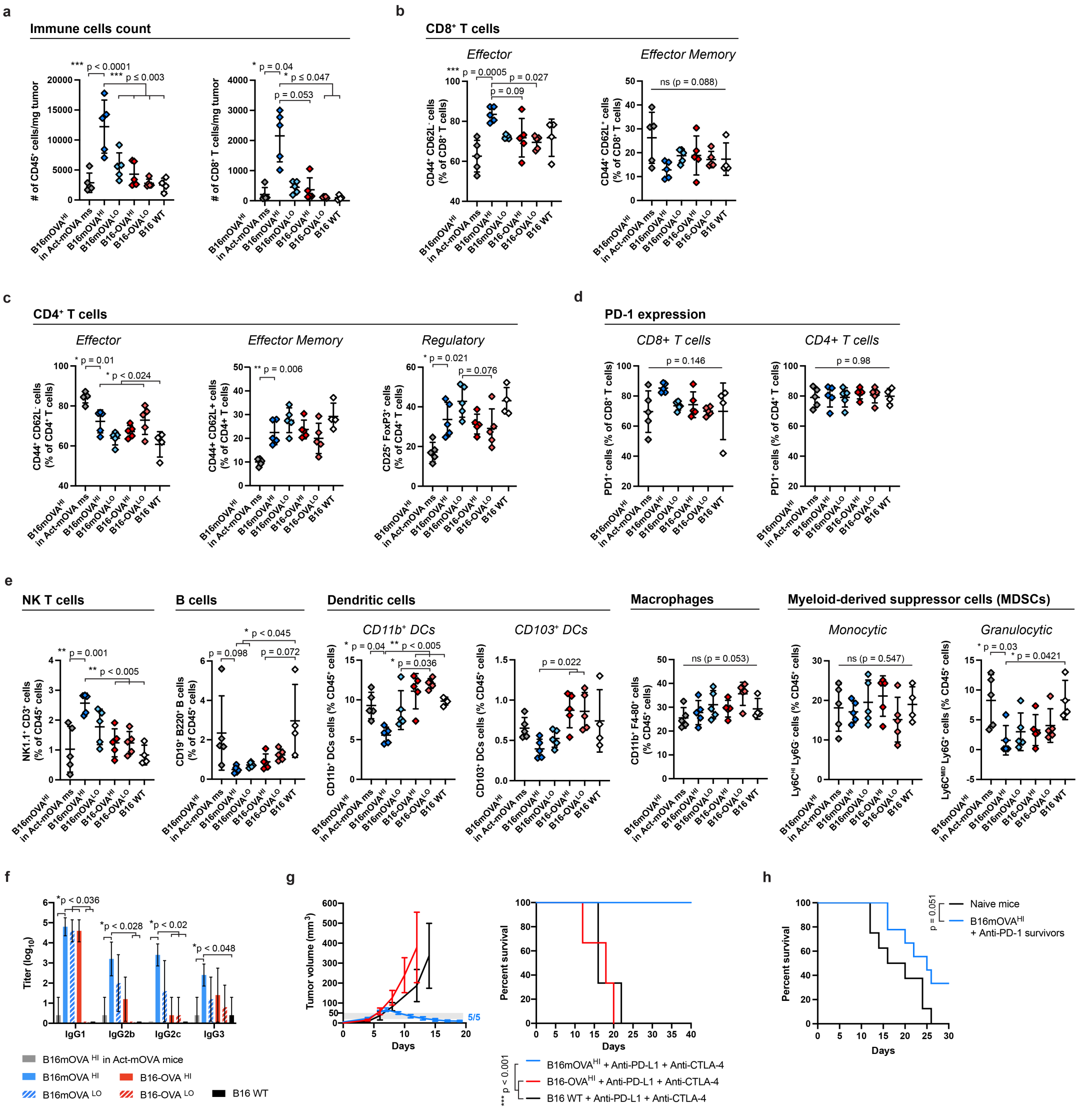


***Supplementary Fig. 4.* Comparison of B16mOVA and B16-OVA melanoma tumor immunogenicity and responsiveness to cancer immunotherapy in mice. a-e,** Flow cytometry analysis of immune cells infiltrated in tumors 10 days post-injection (N≥4, mean ± SD, ANOVA with Tukey's post-test and Brown-Forsythe correction when needed). **a,** Number of $CD45^{+}$ immune cells and $CD8^{+}$ T cells per mg of tumor. **b,** $CD8^{+}$ and **c,** $CD4^{+}$ effector and effector memory T cells subsets in the different tumors. **d,** Proportion of PD-1 expressing $CD8^{+}$ and $CD4^{+}$ T cells. **e,** Proportion of NK T cells, B cells, dendritic cells, macrophages and myeloid-derived suppressor cells relative to the total $CD45^{+}$ immune cell populations. **f,** Titers ($log_{10}$) of the anti-OVA measured per IgG subtype in the plasma of tumor-bearing mice at day 10, which corresponds Fig. 1d. (N≥4, mean ± SD, Kruskal-Wallis with Dunn's post-test per IgG subtype). **g,** Tumor growth and associated survival of OVA-expressing tumor-bearing mice treated with 100 µg of anti-PD-L1 and 100 µg of anti-CTLA-4 injected intraperitoneally when the tumor volume reached 20-50 $mm^3$ (grey thresholds) (N≥3, mean ± SEM, log-rank tests). **h,** Survival of mice re-challenged with B16-F10 WT tumor cells, associated to the tumor growth curves presented in Fig. 1i (N≥8, mean ± SEM, log-rank test).

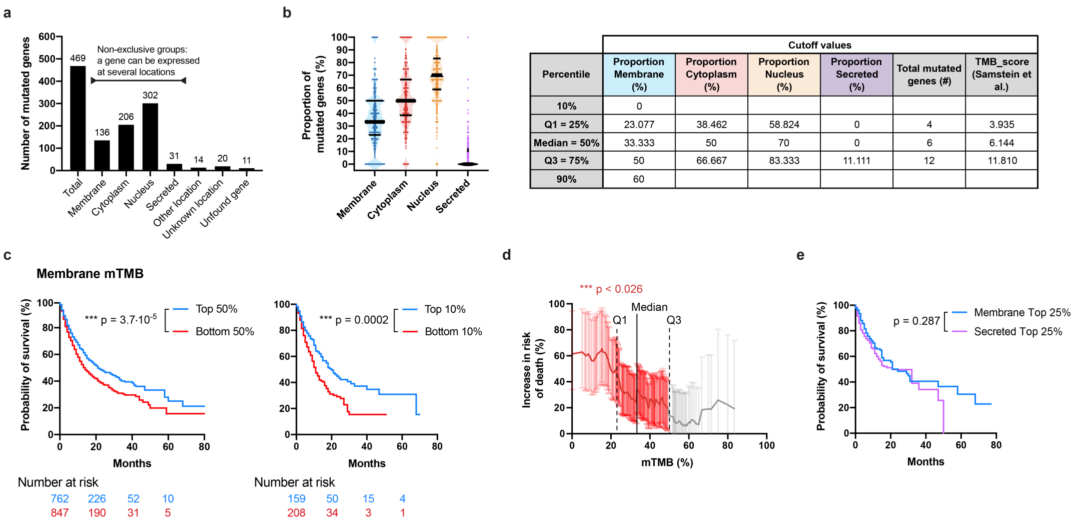


| Percentile | Cutoff values | | | | | |
|---|---|---|---|---|---|---|
| | Proportion Membrane (%) | Proportion Cytoplasm (%) | Proportion Nucleus (%) | Proportion Secreted (%) | Total mutated genes (#) | TMB_score (Samstein et al.) |
| 10% | 0 | | | | | |
| Q1 = 25% | 23.077 | 38.462 | 58.824 | 0 | 4 | 3.935 |
| Median = 50% | 33.333 | 50 | 70 | 0 | 6 | 6.144 |
| Q3 = 75% | 50 | 66.667 | 83.333 | 11.111 | 12 | 11.810 |
| 90% | 60 | | | | | |

***Supplementary Data Fig. 5.*** Analysis of tumor mutated genes' subcellular localizations and their subsequent impact on patient survival. **a,** Number of tumor mutated genes associated with each subcellular location among the 469 genes sequenced by MSK-IMPACT method. **b,** Proportion of tumor mutated genes per subcellular location in patients treated with immunotherapy in the pan-cancer group, and corresponding percentile cutoff values used for the analysis in Fig. 2. **c,** Survival of ICI-treated patients harboring high (Top 50% or 10%) or low (Bottom 50% or 10%) mTMB (log-rank tests). **d,** Increase in risk of death as a function of mTMB in ICI-treated patients in the pan-cancer group. Values are calculated as 100*(HR-1) ± 95% CI with HR the hazard ratio for survival of patients that have less than the depicted proportion as compared to those that have more. As an example, ICI-treated patients that had less than Q1=23% of mutated genes at the membrane had a 50% increased risk of death as compared to those that have more than 23% membrane mutated genes (log-rank tests, red values = p-value ≤ 0.05, grey values = not significant). **e,** Survival of ICI-treated patients with high (Top 25%) mTMB and sTMB (log-rank test).

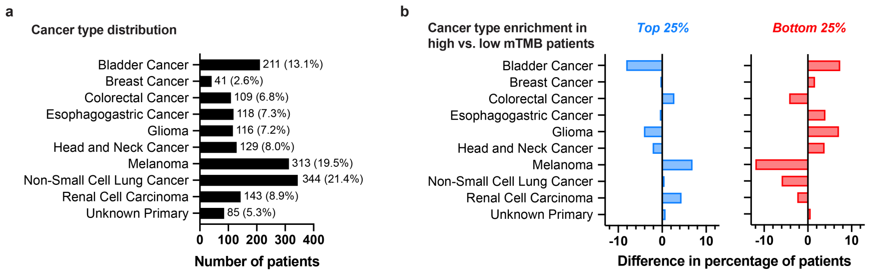


c

Hazard ratio analysis of high vs. low mTMB per cancer type

THRESHOLD USING UNIVERSAL CUTOFF VALUES

| | Hazard ratio | 95% CI upper limit | 95% CI lower limit | p-value | Number of patients Total (bottom vs. top) |
|---|---|---|---|---|---|
| Pan-Cancer | 0.661 | 0.798 | 0.547 | 1.45E-05 | 832 (404 vs. 428) |
| Bladder Cancer | 1.192 | 2.343 | 0.607 | 0.6095 | 104 (84 vs. 21) |
| Breast Cancer | 1.949 | 5.079 | 0.748 | 0.1647 | 26 (17 vs. 9) |
| Colorectal Cancer | 1.015 | 2.732 | 0.377 | 0.9768 | 51 (10 vs. 41) |
| Esophagogastric Cancer | 0.562 | 1.207 | 0.261 | 0.1343 | 75 (46 vs. 29) |
| Glioma | 0.785 | 1.565 | 0.394 | 0.4914 | 71 (58 vs. 13) |
| Head and Neck Cancer | 0.792 | 1.483 | 0.423 | 0.4657 | 73 (48 vs. 25) |
| Melanoma | 0.700 | 1.239 | 0.395 | 0.2183 | 143 (30 vs. 113) |
| Non-Small Cell Lung Cancer | 0.826 | 1.233 | 0.553 | 0.3483 | 156 (62 vs. 94) |
| Renal Cell Carcinoma | 0.351 | 0.690 | 0.178 | 0.0015 | 83 (26 vs. 57) |

THRESHOLD USING 25% TOP vs. 25% BOTTOM

| | Hazard ratio | 95% CI upper limit | 95% CI lower limit | p-value | Number of patients Total (bottom vs. top) |
|---|---|---|---|---|---|
| Pan-Cancer | 0.661 | 0.798 | 0.547 | 1.45E-05 | 832 (404 vs. 428) |
| Bladder Cancer | 0.843 | 1.449 | 0.491 | 0.5369 | 110 (57 vs. 53) |
| Breast Cancer | 1.639 | 4.261 | 0.630 | 0.3061 | 26 (12 vs. 14) |
| Colorectal Cancer | 1.353 | 2.748 | 0.666 | 0.4007 | 69 (28 vs. 41) |
| Esophagogastric Cancer | 0.477 | 1.090 | 0.209 | 0.0731 | 60 (30 vs. 30) |
| Glioma | 1.194 | 2.004 | 0.712 | 0.5013 | 82 (43 vs. 39) |
| Head and Neck Cancer | 0.459 | 0.863 | 0.244 | 0.0135 | 67 (34 vs. 33) |
| Melanoma | 0.827 | 1.295 | 0.528 | 0.4052 | 175 (95 vs. 80) |
| Non-Small Cell Lung Cancer | 0.959 | 1.394 | 0.660 | 0.8277 | 180 (86 vs. 94) |
| Renal Cell Carcinoma | 0.604 | 1.142 | 0.319 | 0.1168 | 102 (45 vs. 57) |

***Supplementary Fig. 6.* Distributions of patients by cancer types and according to their mTMB. a,** Distribution of the ICI-treated patients per cancer type included in the pan-cancer analysis. **b,** Differences in patient distribution per cancer type for the groups with high (Top 25%) or low (Bottom 25%) mTMB, as compared to the distribution of the entire ICI-treated cohort as in panel a. **c,** Values of the HRs, 95% confidence intervals, p-value of the log-rank tests and number of patients used in Fig. 3b, for the universal cutoff and the 25% top vs. bottom strategies.

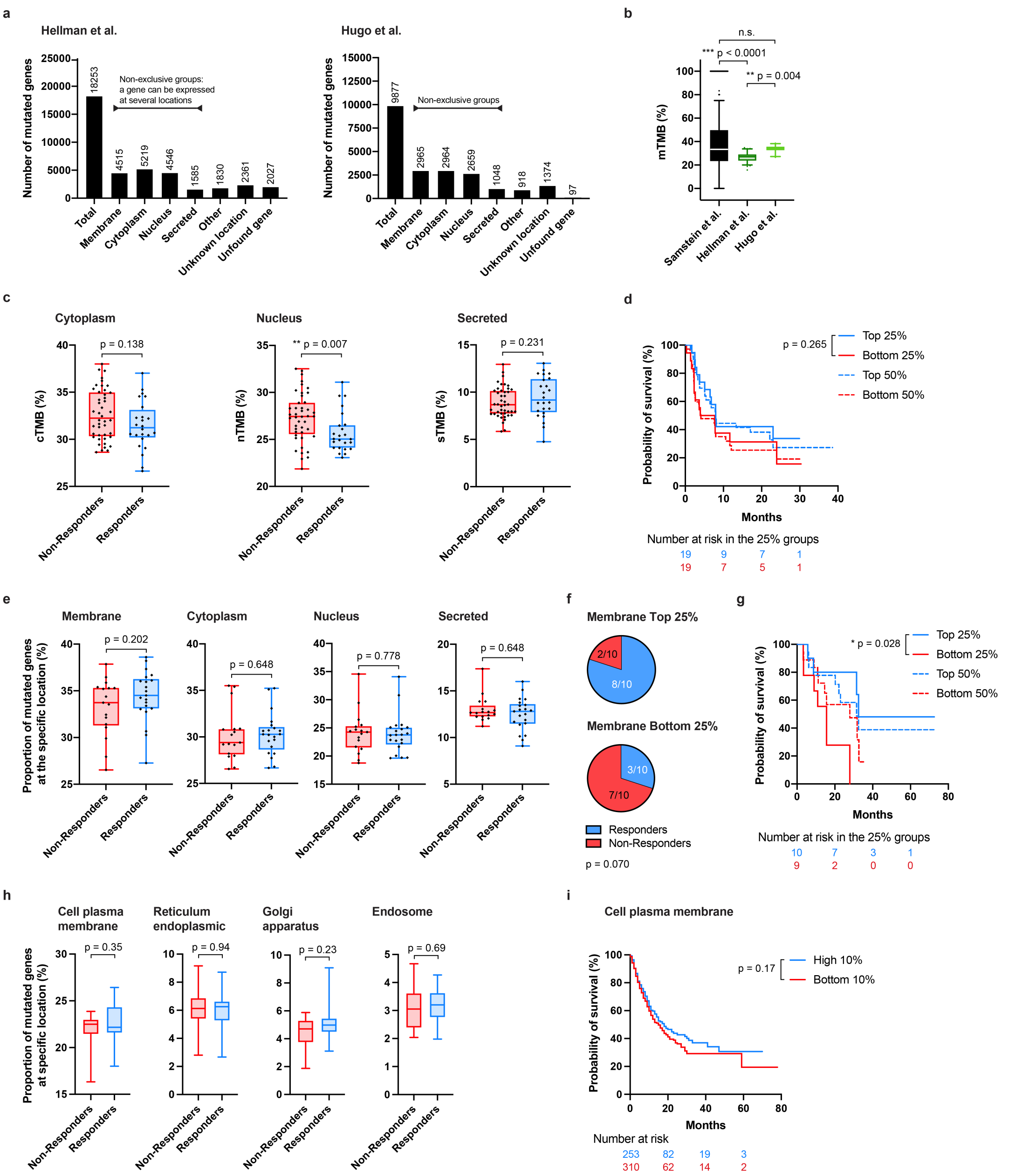


***Supplementary Fig. 7.*** **Response to immunotherapy based on the proportion of mutated genes at specific subcellular localizations.** Patients (N=75) with non-small cell lung cancer were treated with anti-PD-1 + anti-CTLA-4 in the cohort from Hellman *et al.*[8], and patients (N=38) with advanced melanoma cancer were treated with anti-PD-1 in the cohort from Hugo *et al.*[9]. In both studies, tumor mutated genes were sequenced by the WES method. **a,** Number of tumor mutated genes detected across all patients in the Hellman *et al.* and Hugo *et al.* studies, respectively, and their associated subcellular locations. **b,** Comparison of the mTMB found in ICI-treated patient cohorts from the studies by Samstein *et al.*[7], Hellman *et al.*[8] and Hugo *et al.*[9] **c,** Proportion of mutated genes per subcellular location in patients that responded or not to immunotherapy in the Hellman *et al.* cohort (Mann-Whitney test). **d,** Survival of patients with high (Top 25% and 50%) or low (Bottom 25% and 50%) mTMB in the Hellman *et al.*[8] cohort (log-rank test). **e,** Same as in panel c, but with the patient cohort from Hugo *et al.*[9] **f,** Proportion of responders or non-responders to anti-PD-1 among patients that have high (Top 25%) or low (Bottom 25%) mTMB in the Hugo *et al.*[9] cohort (Fisher's exact test). **g,** Same as in panel d, but with the patient cohort from Hugo *et al.*[9]. **h,** Proportion of mutated genes at the cell plasma membrane or in other specific membrane-containing cell organelles in responders and non-responders to immunotherapy from the Hugo *et al.*[9] **i,** Survival of the patients with high (Top 10%) or low (Bottom 10%) proportion of mutated genes expressing proteins at the tumor cell plasma membrane for the pan-cancer groups from the cohort from Samstein *et al.*[7]

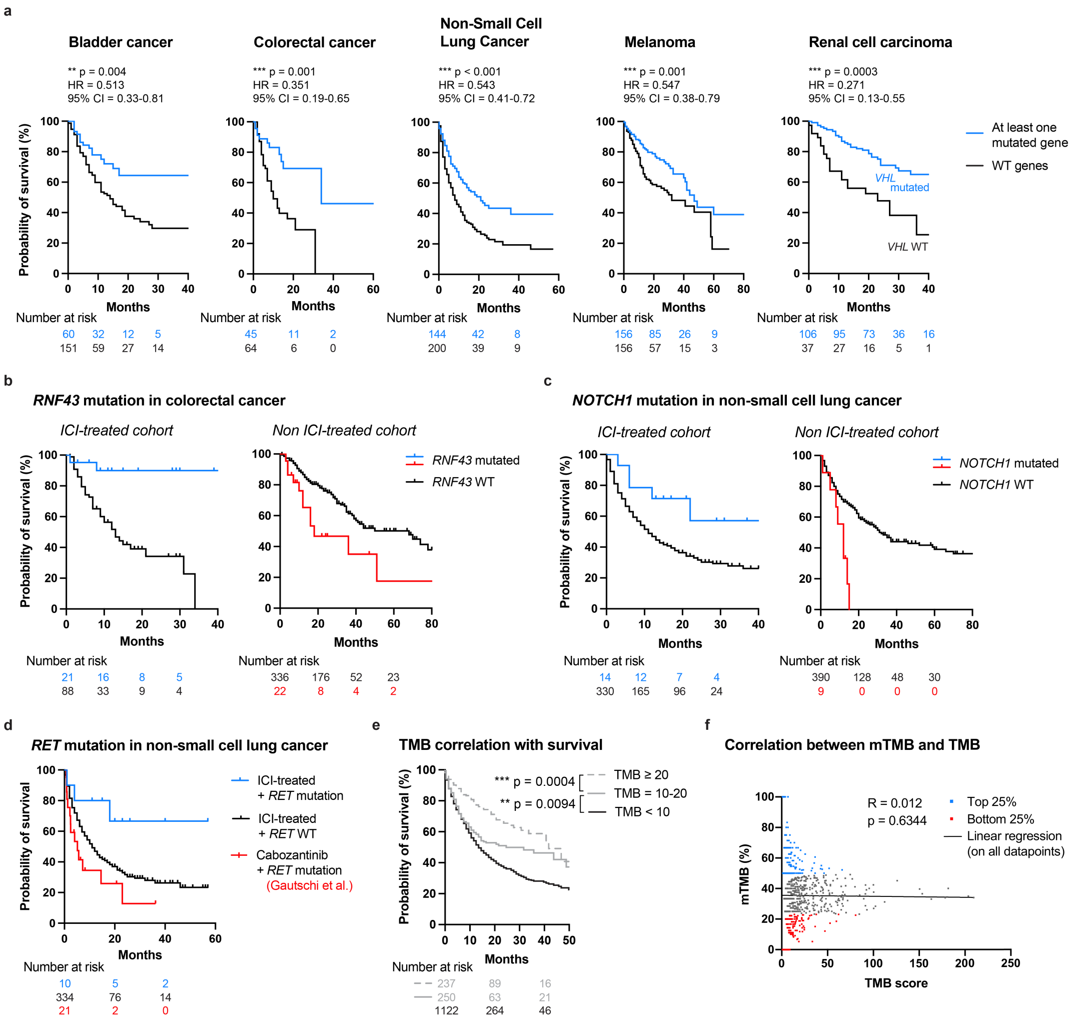


***Supplementary Fig. 8.*** **Potential use of mTMB and specific membrane-associated mutated genes as predictive clinical biomarkers for extended survival upon ICI.** Data analyzed from Samstein *et al.*[7] ICI-treated or non-ICI-treated cohorts. **a,** Survival of patients bearing at least one mutated genes among the cancer-specific list of genes highlighted in blue in Fig. 5a, as compared to patients with no mutated genes among the list. **b,** Survival curves of ICI and non-ICI treated patients bearing *RNF43* mutations in colorectal cancer. **c,** Survival curves of ICI and non-ICI treated patients bearing *NOTCH1* mutations in NSCLC. **d,** Comparison of survival of patients carrying *RET* mutations in NSCLC treated with ICI or with a standard-of-care cabozantinib (data from Gautschi *et al.*[25]). **e,** Survival curves of patients from the pan-cancer group in function of their TMB level (in mut/Mbp). The higher the TMB the longer the survival (log-rank test). f, Correlation between mTMB and total TMB. No correlation was observed between these two parameters (Spearman correlation).
